# Gestational age at birth and body size from infancy through adolescence: findings from analyses of individual data on 253,810 singletons in 16 birth cohort studies

**DOI:** 10.1101/2022.06.01.22275859

**Authors:** Johan L. Vinther, Tim Cadman, Demetris Avraam, Claus T. Ekstrøm, Thorkild I.A. Sørensen, Ahmed Elhakeem, Ana C. Santos, Angela Pinot de Moira, Barbara Heude, Carmen Iñiguez, Costanza Pizzi, Elinor Simons, Ellis Voerman, Eva Corpeleijn, Faryal Zariouh, Gilian Santorelli, Hazel M. Inskip, Henrique Barros, Jennie Carson, Jennifer R. Harris, Johanna L. Nader, Justiina Ronkainen, Katrine Strandberg-Larsen, Loreto SantaMarina, Lucinda Calas, Luise Cederkvist, Maja Popovic, Marie-Aline Charles, Marieke Welten, Martine Vrijheid, Meghan Azad, Padmaja Subbarao, Paul Burton, Puishkumar J. Mandhane, Rae-Chi Huang, Rebecca C. Wilson, Sido Haakma, Sílvia Fernández-Barrés, Stuart Turvey, Susana Santos, Suzanne C. Tough, Sylvain Sebert, Tanis Fenton, Theo J. Moraes, Theodosia Salika, Vincent W.V. Jaddoe, Deborah A. Lawlor, Anne-Marie Nybo Andersen

## Abstract

**Background:** Preterm birth is the leading cause of perinatal morbidity and mortality, and is associated with adverse developmental and long-term health outcomes, including several cardio-metabolic risk factors. However, evidence about the association of preterm birth with later body size derives mainly from studies using birth weight as proxy of prematurity rather than actual length of gestation. We investigated the association of gestational age at birth (GA) with body size from infancy through adolescence.

**Methods and Findings:** We conducted a two-stage Individual Participant Data (IPD) meta-analysis using data from 253,810 mother-children dyads from 16 general population-based cohort studies in Europe, North America and Australasia to estimate the association of GA with standardized Body Mass Index (BMI) and overweight (including obesity) adjusted for confounders. Using a federated analytical tool (DataSHIELD), we fitted linear and logistic regression models in each cohort separately, and combined the regression estimates and standard errors through random-effects study-level meta-analysis providing an overall effect estimate at early infancy (>0.0-0.5 years), late infancy (>0.5-2.0 years), early childhood (>2.0-5.0 years), mid-childhood (>5.0-9.0 years), late childhood (>9.0-14.0 years) and adolescence (>14.0-19.0 years).

GA was positively associated with BMI in the first decade of life with mean differences in BMI z-score (0.01-0.02) per week of increase in GA, however preterm infants reached similar levels of BMI as term infants by adolescence. The association of GA with risk of overweight revealed a similar pattern of results from late infancy through mid-childhood with an increased odds of overweight (OR 1.01-1.02) per week increase in GA. By adolescence, however, GA was slightly negatively associated with risk of overweight (OR 0.98 [95% CI: 0.97:1.00]) per week of increase in GA, and children born very preterm had increased odds of overweight (OR 1.46 [95% CI: 1.03; 2.08]) compared with term.

The findings were consistent across cohorts and sensitivity analyses, despite considerable heterogeneity in cohort characteristics.

**Conclusion:** Higher GA is potentially clinically important for higher BMI in infancy, while the association attenuates consistently with age. By adolescence, preterm children have on average a similar mean BMI to those born term.

## Introduction

Today, one in ten infants are born preterm (<37 completed weeks’ gestation) with an increased risk of perinatal mortality and morbidity that may persist and develop over the life-course (1–3). Global estimates show an increase in preterm birth between 2000 and 2014, but the proportions vary between countries (4).

Previous systematic reviews and meta-analyses (5–7) have reported an association of gestational age at birth (GA) with conventional cardio-metabolic risk factors, including increased blood pressure, impaired glucose regulation, and insulin resistance in those born preterm (8–11). An infant born preterm adapts to extrauterine conditions entering a phase of growth that possibly expresses a mismatch with the environment outside utero leading to alterations in body composition (12–17). It has been hypothesized that these changes increase susceptibility to being overweight for preterm birth through various pathways and mechanisms, including catch-up weight (16, 18–22). However, later body size in preterm cohorts is not well characterized and most studies define populations by birth weight rather than actual length of gestation (17, 23). It is recognized that determinants and consequences of gestational duration are quite different from those of foetal growth (23), and that birth weight reflects both gestational duration and foetal growth (24), hence being at potential intermediate variable on the causal pathway (25).

Studies have shown that infants born extremely (23-27 weeks gestation) and very preterm (28-31 weeks gestation) typically experience postnatal growth failure followed by catch-up weight and length gain within the first two years of life (20). Growth in preterm children remains different from that of full term peers through childhood and into school age (26–32). However, studies on growth in preterm cohorts across key stages of growth development (33) and at more advanced gestational age are scarce (10, 20, 34). Several methodological considerations and sample characteristics complicate the interpretation and comparability of findings on the relationship between GA with later body size (5, 6, 35–37). This includes differences in study design; using birth weight as a proxy for GA; sample size; age at outcome; conditions under which variables are examined; type of statistical analysis; and availability of confounders.

In this study, we use the novel approach and unique opportunity of federated analysis of individual participant data in a secure manner provided by the EU Child Cohort Network (38, 39), an international network of European and Australasian birth cohort data. We base our study on 16 cohorts and 253,810 mother-child dyads, which enables us to extend previous research by including information on repeated body size measures during a long follow-up across a wide range of GA, and overcome the methodological limitations identified above.

The overall aim of this study was to determine the association between GA (completed weeks and clinical categories) and, respectively, body mass index (BMI) and overweight (including obesity) from infancy through adolescence in birth cohort studies representing diverse contexts.

## Methods

### Inclusion criteria and participating cohorts

In December 2019, we invited pregnancy and birth cohort studies within the EU Child Cohort Network from the LifeCycle and the EUCAN-Connect consortia (38–40). Cohorts were eligible for inclusion if they had information for live-born singletons on GA and at least one offspring measurement of BMI in one of six age-periods: early infancy (>0.0-0.5 years), late infancy (>0.5-2.0 years), early childhood (>2.0-5.0 years), mid-childhood (>5.0-9.0 years), late childhood (>9.0-14.0 years), adolescence (>14.0-19.0 years).

The following 16 cohorts participated in the study: Avon Longitudinal Study of Parents and Children, UK (*ALSPAC*) (n=10,452) (41), All Our Families, Canada (*AOF*) (n=2,263) (42), Born in Bradford, UK (*BIB*) (n=13,097) (43), CHILD Cohort Study (*CHILD*) (n=2,984) (44), Danish National Birth Cohort, Denmark (*DNBC*) (n=81,117) (45), The EDEN mother-child cohort on the prenatal and postnatal determinants of child health and development, France (*EDEN*) (n=1,765) (46), French Longitudinal Study of Children, France (*ELFE*) (n=15,506) (47), The Generation 21 Birth Cohort, Portugal (*G21*) (n=6,439) (48), The GECKO Drenthe Cohort, The Netherlands (*GECKO*) (n=2,768) (49), The Generation R Study, The Netherlands (*GEN-R*) (n=8,641) (50), The Environment and Childhood Project, Spain (*INMA*) (n=1,936) (51), The Norwegian Mother, Father and Child Study, Norway (*MoBa*) (n=86,553) (52), The Northern Finland Birth Cohort 1986, Finland (*NFBC1986*) (n=8,325) (53–55), The NINFEA (Nasita e INFanzia: gli Effetti dell’Ambiente) birth cohort study, Italy (*NINFEA*) (n=6,515) (56), The Raine Study, Australia (*The Raine Study*) (n=2,443) (57), and The Southampton Women Survey, UK (*SWS*) (n=3,007) (58).

### Data Access and Federated Analysis on DataSHIELD

In this study, we used pseudonymized data stored on local secure data servers in their original location (59–62), and harmonised according to protocols in the EU Child Cohort Network (39). Cohort-specific description about methods for ascertaining and defining variables are documented in the EU Child Cohort Network catalogue (https://data-catalogue.molgenis-cloud.org/catalogue/catalogue/#/) and the Maelstrom Catalogue (http://maelstrom-research.org) for studies in LifeCycle and EUCAN-Connect, respectively. Data were analysed remotely through the open-source software, DataSHIELD, which allows both federated study level and Individual Participant Data (IPD) meta-analysis with active disclosure protection (63–65). Fourteen cohorts gave permission to analyse their data via DataSHIELD, and two cohorts (AOF, CHILD) via data transfer agreements.

### Gestational age at birth

Information on GA (in days) was available as harmonised IPD with source of delivery information obtained from medical records in the majority of cohorts (S1 Table, S1 Text). GA was rounded to completed weeks, and further categorized into five groups (66): 28-33 weeks (very preterm), 34-36 weeks (late preterm), 37-38 weeks (early term), 39-41 weeks (full term) and 42-43 weeks (post term).

### Offspring BMI and overweight and obesity

Information on height (cm) and weight (kg) was available as harmonised IPD measured in either a clinical setting, or selfreported by parents or index child (Table S1). BMI was calculated as weight (kg) / (height (m))^2^ (67), and sex-and-age specific BMI z-scores were calculated per month using external WHO standards (68) and references (69). We defined overweight (including obesity) following WHO cut-offs, separately for children <5 years (>2 standard deviations above WHO Child Growth Standard median) and ≥5 years (>1 standard deviation above WHO Growth Reference median). In several cohorts (ALSPAC, BIB, DNBC, GEN-R, INMA, NINFEA, NFBC1986, the Raine Study, SWS), multiple measurements of BMI were available for the same child within one or more of the six age-groups. In such cases, the latest measurement was chosen.

### Confounders

Confounders were selected a priori as factors that were known or plausible causes of variation in GA and subsequent body size with a directed acyclic graph used in discussions to select the final set of confounders (S2 Table). The resulting confounders were: maternal education (ISCED-2011/97, low/medium/high) (S1 Text) (70), maternal height (continuous, m), maternal pre-pregnancy BMI (continuous, kg/m^2^), maternal smoking during pregnancy (yes/no), maternal age at child’s birth (continuous, years); gestational diabetes (yes/no); gestational hypertension (yes/no); pre-eclampsia (yes/no), maternal ethnic background (western/non-western/mixed) (S1 Text), and parity (nulliparous/parous). For the objective of this study, we did not include birth weight as it may distort interpretation of the results being an intermediate variable on the causal pathway (25).

### Statistical Analysis

Distributions of GA at birth, body size measures and confounders were obtained for each cohort separately, and for all cohorts combined.

We conducted a two-stage meta-analysis to estimate associations between GA with BMI and overweight, adjusted for confounders. We fitted a linear regression model to examine the associations of GA in weeks and in clinical categories with BMI z-scores. Models were fitted in each cohort separately, and cohort-specific coefficients and standard errors were combined and assigned weights using random-effects model to attain overall effect estimates (71, 72). The analyses were performed separately for the six age-groups (>0.0-0.5 years, >0.5-2.0 years, >2.0-5.0 years, >5.0-9.0 years, >9.0-14.0 years, >14.0-19.0 years). To examine the associations between GA in weeks and in clinical categories and odds of overweight (compared with normal weight) we used a binomial logistic regression model.

The main results are those from regression analyses adjusted for the maximum set of baseline confounders available within each cohort. Models were adjusted for maternal age at child’s birth, height, education, pre-pregnancy BMI, and parity in all cohorts. Models were additionally adjusted for maternal ethnic background, gestational hypertension, gestational diabetes, and preeclampsia in cohorts where these were available (Table 1).

**Table 1.**
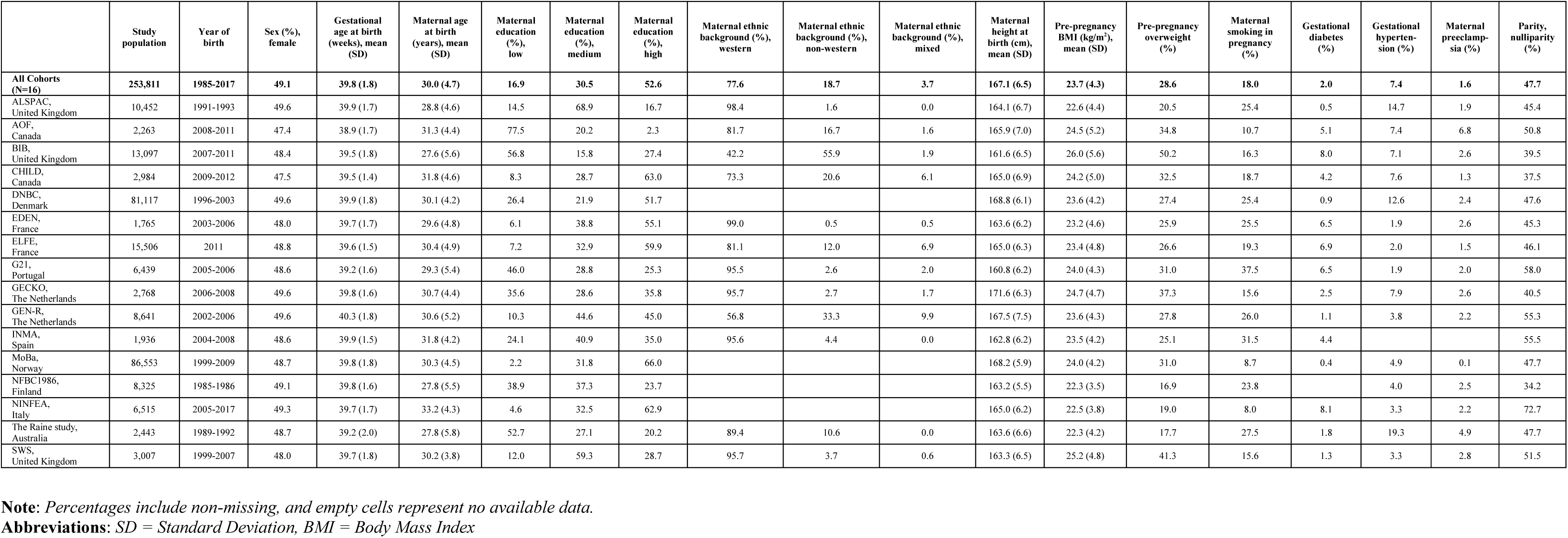
Baseline characteristics of study participants in the 16 participating cohorts.

Results are presented with 95% confidence intervals, and I^2^ statistics (73). We examined between-study heterogeneity by meta-regression in meta-analyses with considerable heterogeneity reflected by either I^2^ > 75% or I^2^ ∼ 75% with effect estimates in different directions. The meta-regressions were conducted to determine which study characteristics were independently associated with between-study heterogeneity. In addition, we undertook ‘Leave-one-out’-analysis for cross-validation to explore the influence of each study on the overall estimate (74), while sub-group analysis with sex (boys vs. girls), maternal education (high vs. low/medium), and maternal smoking in pregnancy (no vs. yes) was performed to measure the robustness of our findings.

Statistical analyses were performed using DataSHIELD (dsBaseClient v6.1.0, https://github.com/datashield/dsBaseClient/releases) and the Statistical Software R (v4.1) (75). We used the ds.getWGSR and ds.glmSLMA functions and the dh.makeStrata function from the ds.Helper-package in addition to the rma-package v3.0.2 (76). Forest plots were created using Excel 2016.

### Ethical approval

Cohort-specific ethical approvals and consent from participants are listed in Supplementary Information (S3-S4 Appendix).

## Results

### Descriptive statistics

A total of 253,810 mother-child dyads in 16 cohort studies from 11 countries had information on GA and at least one measurement of offspring BMI.

Descriptive information including characteristics of GA at birth, body size measures and covariates for study participants are displayed separately for each cohort and for the cohorts combined in Table 1-3.

**Table 2.** Distribution of gestational age groups in the 16 participating cohorts.

|  | Gestational age at birth (completed weeks) |  |  |  |  |
| --- | --- | --- | --- | --- | --- |
|  | Very preterm<br>28-33 weeks (%) | Late preterm,<br>34-36 weeks (%) | Early term,<br>37-38 weeks (%) | Full term,<br>39-41 weeks (%) | Post term,<br>42-43 weeks (%) |
| **All Cohorts**<br>**(N=16)** | **1.2** | **4.3** | **17.8** | **69.9** | **6.7** |
| ALSPAC,<br>United Kingdom | 1.2 | 4.2 | 17.0 | 69.9 | 7.7 |
| AOF,<br>Canada | 1.3 | 5.0 | 26.1 | 67.4 | 0.2 |
| BIB,<br>United Kingdom | 1.4 | 4.6 | 22.5 | 70.2 | 1.3 |
| CHILD,<br>Canada |  | 4.2 | 23.5 | 71.5 | 0.8 |
| DNBC,<br>Denmark | 1.3 | 4.3 | 16.4 | 69.5 | 8.6 |
| EDEN,<br>France | 1.4 | 4.0 | 18.9 | 73.6 | 2.0 |
| ELFE,<br>France | 0.5 | 4.9 | 20.6 | 73.5 | 0.5 |
| G21,<br>Portugal | 1.4 | 5.8 | 31.6 | 61.1 | 0.2 |
| GECKO,<br>The Netherlands | 0.7 | 4.3 | 19.9 | 70.4 | 4.7 |
| GEN-R,<br>The Netherlands | 1.0 | 3.5 | 11.8 | 68.4 | 15.4 |
| INMA,<br>Spain | 0.4 | 2.7 | 19.3 | 71.8 | 5.8 |
| MoBa,<br>Norway | 1.4 | 4.4 | 16.7 | 70.0 | 7.6 |
| NFBC1986,<br>Finland | 1.3 | 3.5 | 17.5 | 73.9 | 3.9 |
| NINFEA,<br>Italy | 1.1 | 4.5 | 21.2 | 69.2 | 4.0 |
| The Raine study,<br>Australia | 2.1 | 5.4 | 20.8 | 65.9 | 5.8 |
| SWS<br>United Kingdom | 1.5 | 4.4 | 17.3 | 72.4 | 4.5 |

**Note:** Empty cells represent no available data

**Table 3.**
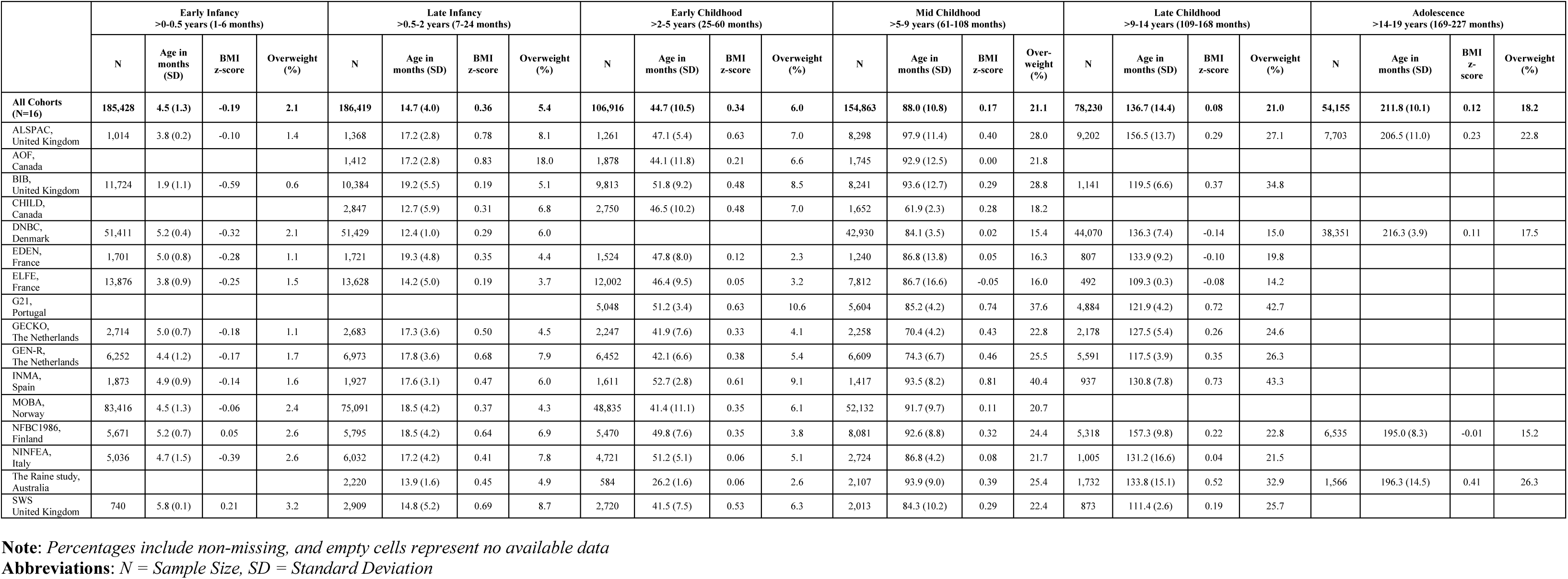
Distribution and age of body size measurements in the 16 participating cohorts.

There were distinct differences in the cohort-specific sample sizes (n=1,765-86,553), distributions of maternal education (range: 2.2% to 77.5% for low), maternal ethnicity (range: 42.2% to 99.0% for Western; 0.5% to 55.9% for non-Western; 0.0% to 9.9% for mixed); maternal pre-pregnancy overweight (range: 16.9% to 50.2%), gestational hypertension (range: 1.9% to 19.3%), and parity (range for nulliparous: 34.2% to 72.7%) (Table 1).

The mean gestational age was 39.8 weeks and overall 5.5% were born preterm (range: 3.1% to 7.5%), 17.8 % (range: 11.8% to 31.6%) were born early term, 69.9% were born full term (range 61.1% to 73.6%), and 6.7% (range: 0.2% to 15.4%) were born post term (Table 2). The majority of the cohorts had study participants included for analysis in all five categories of GA, except CHILD (34-43 weeks gestation).

From infancy to age 19 years, 711,856 measurements of BMI were available for 253,810 children. The number of cohorts and participants with data on BMI and overweight varied across the six age-bands with most cohorts and participants in infancy and mid-childhood and fewest in adolescence, where four cohorts (ALSPAC, DNBC, NFBC1986, the Raine Study) contributed with data on 36,895 individuals. The proportion of children classified as overweight also varied between cohorts, and across age-bands due to different cut-offs used for children < five years and in children ≥ five years (Table 3).

The percentage of missing values for baseline characteristics is presented in Supplementary Information (S4 Table).

### Gestational age at birth and BMI z-scores

The overall estimates for the associations of GA in completed weeks and clinical categories with BMI z-score are displayed in Fig. 1 and Fig. 2.

**Fig 1.**
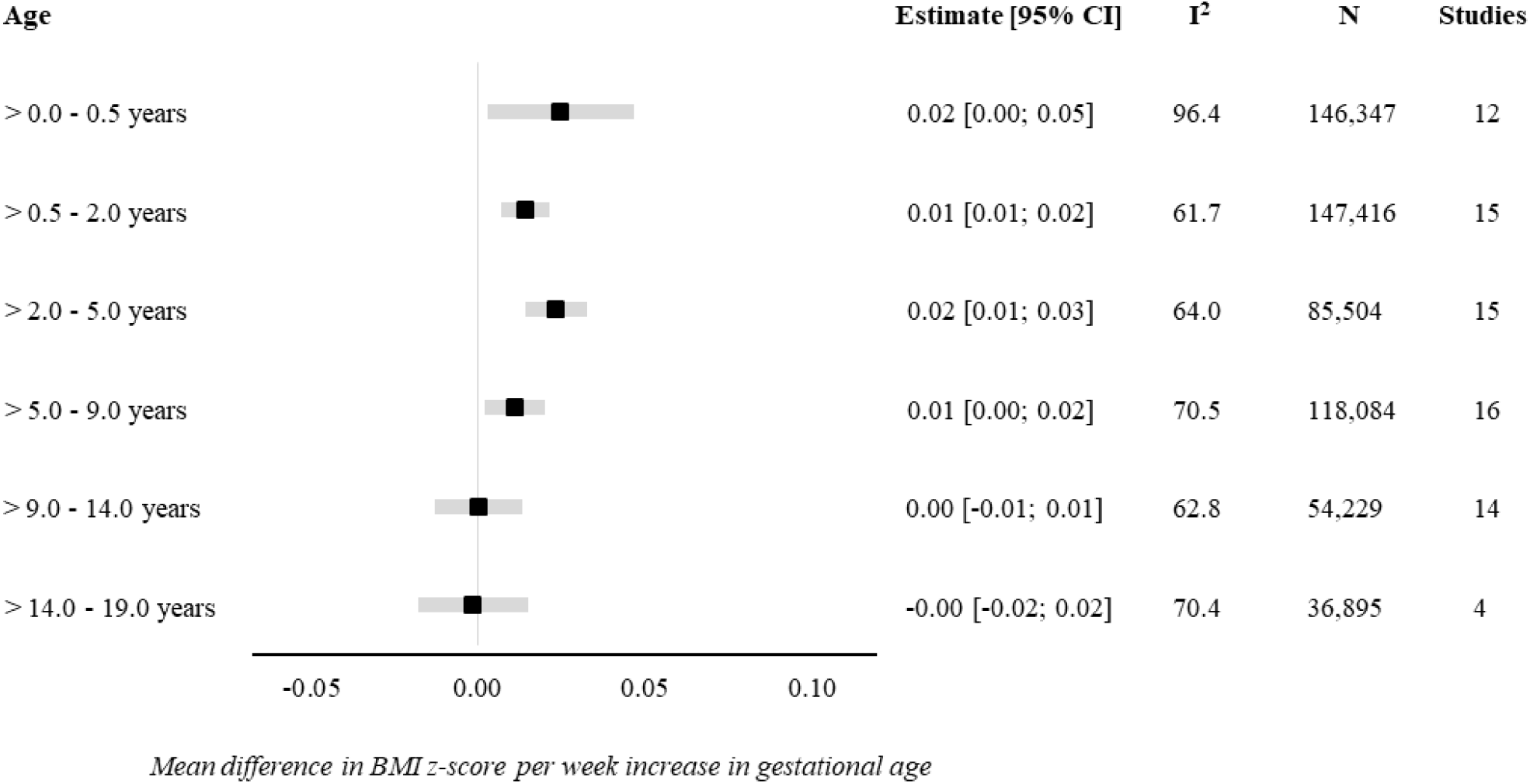
Forest plot of associations between gestational age at birth (completed weeks) and BMI z-score. Values are overall estimates from IPD meta-analyses of the study-specific linear regression model estimates, where cohorts were assigned weights under the random-effects model. Overall estimates reflect a mean difference in BMI z-scores per week increase in gestational at birth in early infancy (>0-0.5 years), late infancy (>0.5-2.0 years), early childhood (>2-5 years), mid childhood (>5-9 years), late childhood (>9-14 years), and adolescence (>14-19 years). Models are adjusted for sex of child, maternal age at child’s birth, maternal education, maternal height, maternal pre-pregnancy BMI, maternal smoking during pregnancy, parity, maternal ethnic background, gestational diabetes, gestational hypertension, pre-eclampsia. Specific variables were not available in few cohorts (see Table 1), hence not adjusted for.

**Fig 2.**
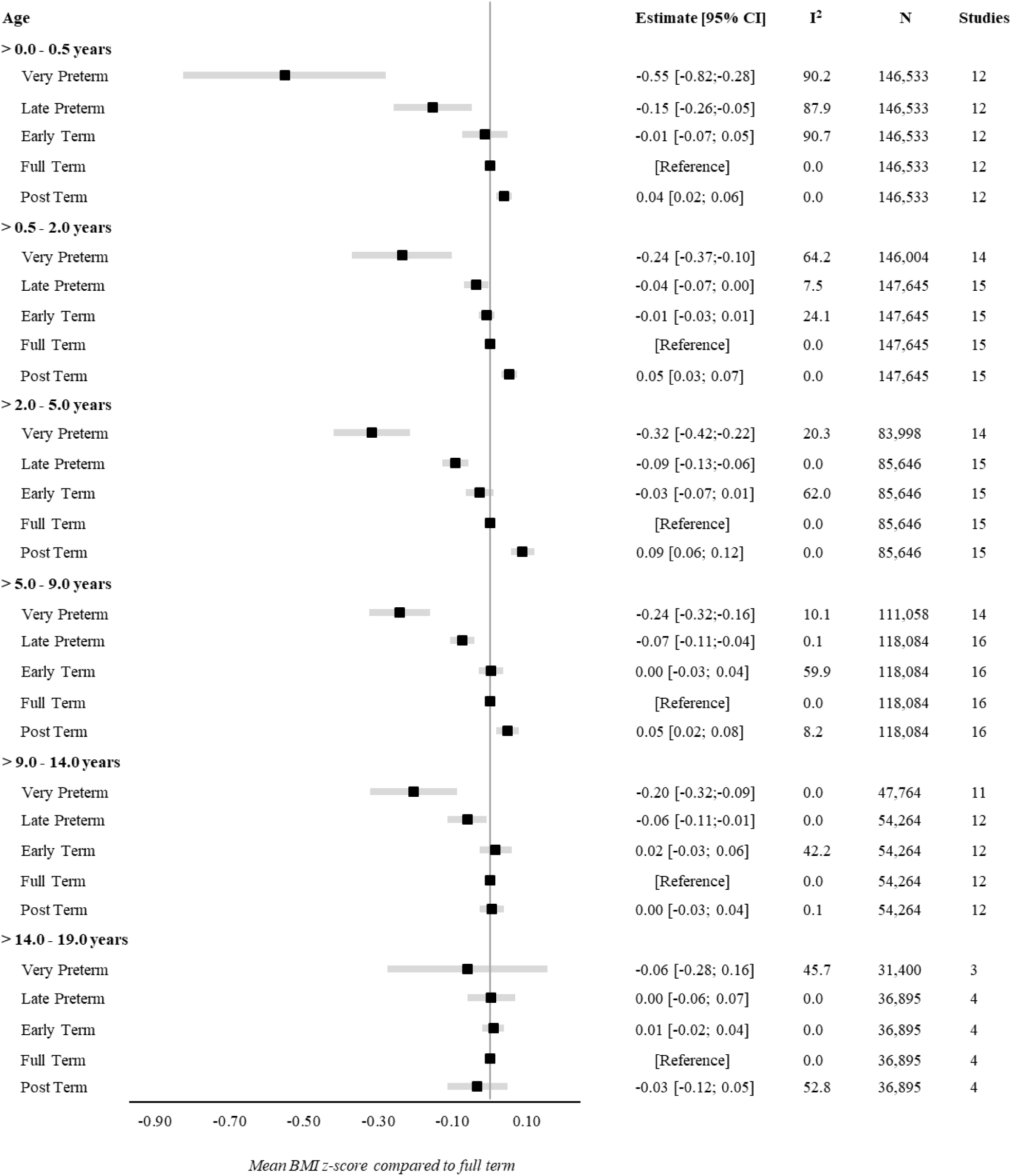
Forest plot of associations between gestational age at birth (clinical categories) and BMI z-score. Values are overall estimates from IPD meta-analyses of the study-specific linear regression model estimates, where cohorts were assigned weights under the random-effects model. Overall estimates reflect a mean BMI z-scores compared to full term (reference category) in early infancy (>0-0.5 years), late infancy (>0.5-2.0 years), early childhood (>2-5 years), mid childhood (>5-9 years), late childhood (>9-14 years), and adolescence (>14-19 years). Models are adjusted for sex of child, maternal age at child’s birth, maternal education, maternal height, maternal pre-pregnancy BMI, maternal smoking during pregnancy, parity, maternal ethnic background, gestational diabetes, gestational hypertension, pre-eclampsia. Specific variables were not available in few cohorts (see Table 1), hence not adjusted for.

There was a positive association of GA with BMI in early infancy (>0.0-0.5 years): 0.02 SD per week increase in GA [95% CI: 0.00, 0.05] was associated with a BMI z-score of -0.55 [95% CI: -0.82, -0.28] for very preterm and -0.15 [95% CI: - 0.26, -0.05] for late preterm compared to full term. Results attenuated through childhood and continuing to decrease to zero by adolescence (0.00 [95% CI: -0.02, 0.02]) with no difference in BMI z-score between preterm and full term peers.

Between-study heterogeneity was examined through meta-regression in four age-bands (>0.0-0.5 years, >5.0-9.0 years, >9.0-14.0 years, >14.0-19.0 years) having considerable heterogeneity, with largest I^2^-statistics (96.4%) in early infancy (S4 Table). We examined age at measurement, child sex, maternal education and maternal smoking in pregnancy as between-study characteristics. The meta-regression found age at measurement to be significantly associated with heterogeneity in early infancy (β=-0.03, se=0.008, p<0.01); maternal education in late childhood (β=0.001, se=0.001, p=0.05), and both maternal education (β=0.001, se=0.001, p<0.01) and smoking in pregnancy (β=0.01, se=0.003, p=0.01) in adolescence.

The ‘Leave-one-out’ analyses gave similar overall effect estimates in all age-bands and did not change between-study heterogeneity markedly (S2 Fig.), however in adolescence leaving out ALSPAC changed the I^2^ from 70.4% to 0.4% (S2 Fig. F). Sub-group analyses were consistent with the main findings across sex (S3 Fig.), maternal educational level (S4 Fig.), and pregnancy smoking-status (S5 Fig.).

### Gestational age at birth and overweight

The overall estimates for the associations of GA in completed weeks and clinical categories with odds of overweight are displayed in Fig. 3 and Fig. 4.

**Fig 3.**
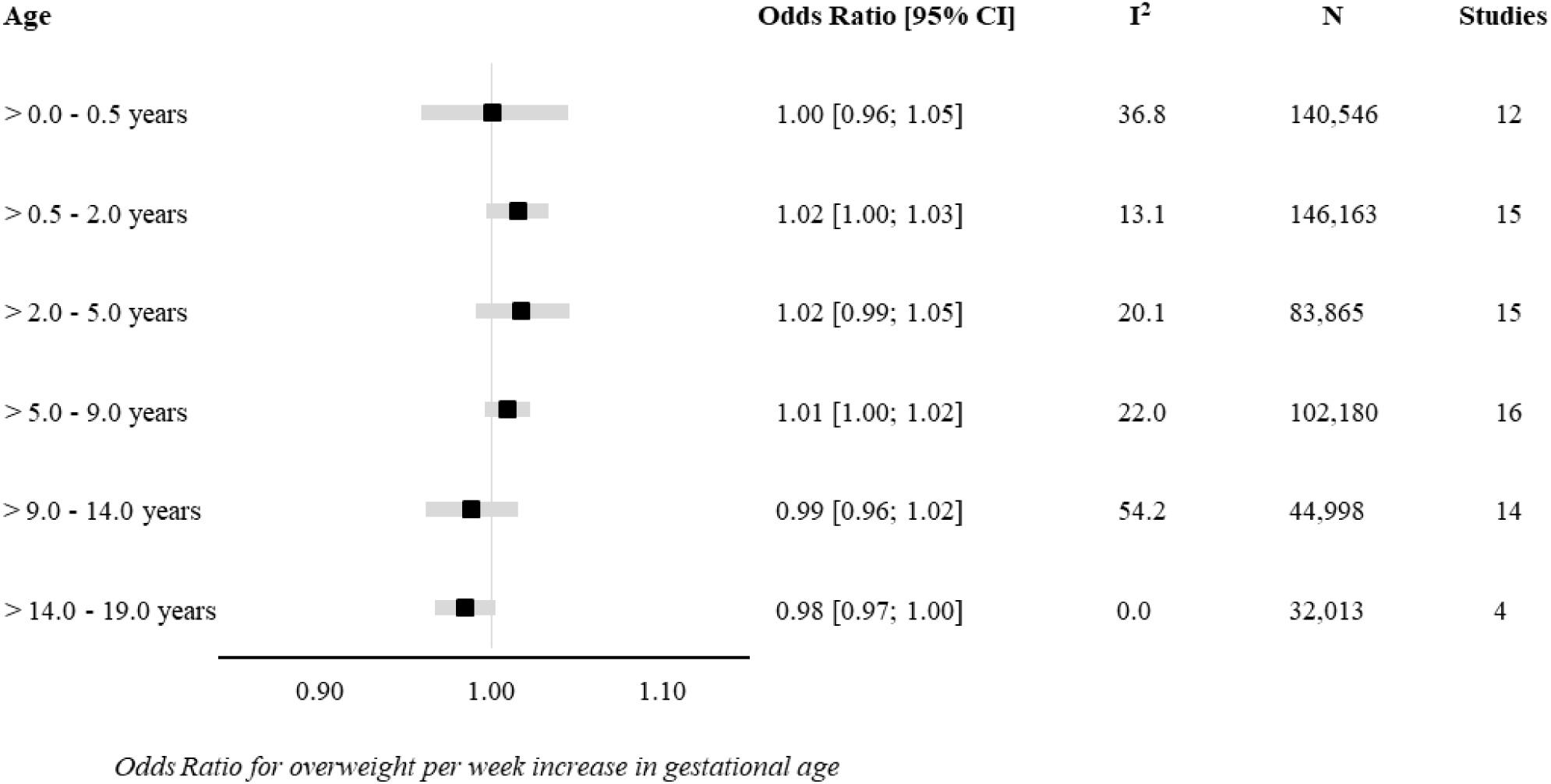
Forest plot of associations between gestational age at birth (completed weeks) and odds of overweight. Values are overall estimates from IPD meta-analyses of the study-specific logistic regression model estimates, where cohorts were assigned weights under the random-effects model. Overall estimates reflect odds ratio for overweight per week increase in gestational at birth in early infancy (>0-0.5 years), late infancy (>0.5-2.0 years), early childhood (>2-5 years), mid childhood (>5-9 years), late childhood (>9-14 years), and adolescence (>14-19 years). Models are adjusted for sex of child, maternal age at child’s birth, maternal education, maternal height, maternal pre-pregnancy BMI, maternal smoking during pregnancy, parity, maternal ethnic background, gestational diabetes, gestational hypertension, pre-eclampsia. Specific variables were not available in few cohorts (see Table 1), hence not adjusted for.

**Fig 4.**
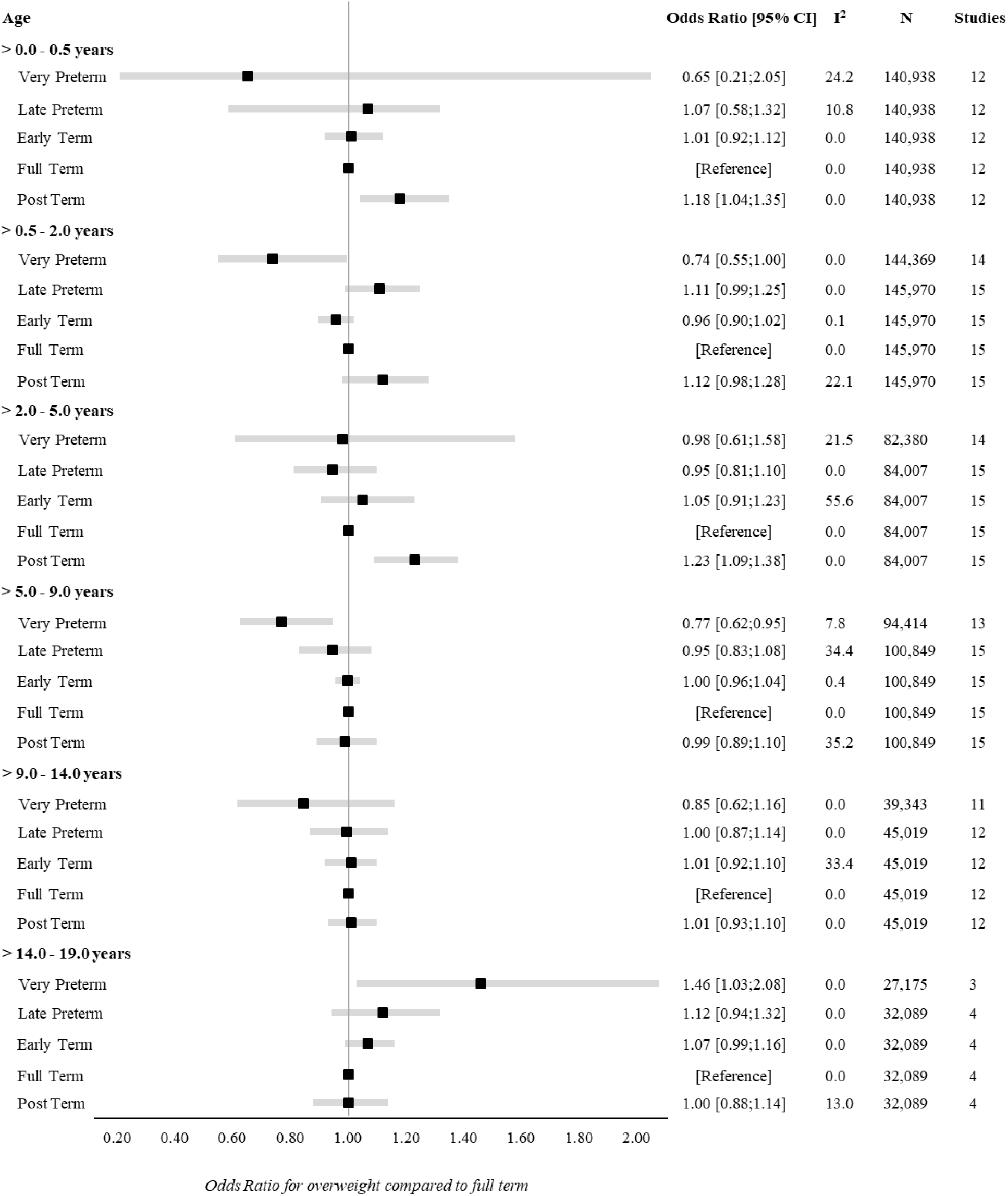
Forest plot of associations between gestational age at birth (clinical categories) and odds of over-weight. Values are overall estimates from IPD meta-analyses of the study-specific logistic regression model estimates, where cohorts were assigned weights under the random-effects model. Overall estimates reflect odds ratio for overweight compared to full term (reference category) in early infancy (>0-0.5 years), late infancy (>0.5-2.0 years), early childhood (>2-5 years), mid childhood (>5-9 years), late childhood (>9-14 years), and adolescence (>14-19 years). Models are adjusted for sex of child, maternal age at child’s birth, maternal education, maternal height, maternal pre-pregnancy BMI, maternal smoking during pregnancy, parity, maternal ethnic background, gestational diabetes, gestational hypertension, pre-eclampsia. Specific variables were not available in few cohorts (see Table 1), hence not adjusted for.

There was a positive association of GA with odds of overweight (OR 1.02 per week increase in GA) in late infancy [95% CI: 1.00, 1.03] and early childhood [95% CI: 0.99, 1.05]. Results attenuated through childhood and continued to decrease to below one by late childhood. In adolescence (>14.0-19.0 years), there was a negative association of GA with odds of over-weight with very preterm having a significantly increased risk of overweight (OR 1.46 [95% CI: 1.03, 2.08] compared with full term peers.

None of the five age-bands had considerable between-study heterogeneity (I^2^ < 55%); hence, we did not perform meta-regression for the associations of GA with odds of overweight. The ‘Leave-one-out’ analyses were consistent with the main findings without changing the overall effect estimate in any of the age-bands or any notable changes in the between-study heterogeneity (S7 Fig.). The subgroup-analysis showed no difference in the associations of GA with odds of overweight for sex (S8 Fig.), maternal educational level (S9 Fig.) or pregnancy smoking-status (S10 Fig.).

## Discussion

In this two-stage meta-analysis using individual participant data on 253,810 live-born singletons from 16 birth cohorts, we found a potentially important association in early infancy between GA and BMI, and in adolescence for the association of GA with odds of overweight. Difference in BMI z-score between categories of GA attenuated markedly after infancy throughout adolescence. A similar trend was observed for the association of GA with odds of overweight; however, by adolescence increased odds of overweight was observed in very preterm compared with full term peers. Despite heterogeneity in cohort characteristics, our main findings were consistent across cohorts and the supplementary analyses showed associations to be robust.

### Interpretation of main findings

Previous studies (26–28, 77) and a meta-analysis (35) have shown consistent results for the association of GA with BMI in childhood with lower BMI in preterm children compared with full term peers, although several methodological issues should be taken into account when interpreting these findings. In contrast, surprisingly few studies have examined the association of GA with later overweight, particularly in childhood.

Existing evidence for the association of GA and BMI rely on small sample sizes from different countries; different GA-categorization and reference group; and variations in use of BMI indices (z-scores or natural units, external or internal reference, IOTF or WHO reference). In our study, we observed a positive overall estimate for the association between GA and BMI in early infancy through mid-childhood with lower BMI z-score in very and late preterm compared to full term peers. In addition, we found age to be the main driver of between-study heterogeneity in early infancy suggesting that GA has a potentially important association in infancy. Our findings are in line with descriptive results from Australia (26) and Sweden (27). In an Australian cohort the authors reported lower BMI z-scores among 225 extremely preterm compared with 253 term controls at both 2 and 5 years, and researchers in Sweden found a lower mean BMI z-scores at 2 and 5 years among 152 Swedish children born between 32-37 weeks compared with a large reference population. Our study showed a weaker association that most likely is explained by adjustment for confounders, but also methodological differences.

Our analyses revealed that the overall associations between GA and BMI attenuated in mid- and late childhood, but very and late preterm children remained at a lower BMI compared to their full term counterparts. Similar associations were reported in studies from Brazil and United Kingdom on 203 and 497 children aged 6 and 8-12 years, respectively (28, 77). Moreover, the study from Brazil compared BMI z-scores in 350 very (<33 weeks) and late preterm (34-36 weeks) aged 18 years with 3,027 early and full term peers using the WHO Growth Reference. In line with our findings, children born preterm in Brazil reached similar BMI levels of term counterparts by adolescence supporting that the overall association attenuates through childhood (20, 26).

A rapid phase of growth has been proposed to evolve into increased susceptibility of later overweight (21, 78–80), but only few studies have examined the relationship between GA and later overweight in childhood or adulthood (10, 29, 30, 81). The overall effect estimates from our main analysis showed a weak association between GA and overweight from early infancy through mid-childhood with only very preterm in mid-childhood being at lower odds of overweight than full term peers. In contrast, a cohort study from Chile based on 153,635 children aged 6-8 years reported that term children are a lower risk of overweight (OR 0.84 [95% CI: 0.79, 0.88]) than preterm peers (reference group, (≤37 weeks) (29). However, as highlighted by the authors, a major limitation of their study was the lack of information on obstetric maternal characteristics and maternal pre-pregnancy BMI.

In accordance with a cohort study from the United Kingdom on 11,765 children aged 11 years (30), we found no difference in odds of overweight between preterm and full term children in late childhood (>9.0-14.0 years).

Our study extends previous research by examining the association between GA and overweight in adolescence, and across key stages of growth development throughout childhood. Moreover, our study design and large sample size enables an examination of odds of overweight in preterm adolescents, and provides insights about this association across a wide range of GA. This distinction between degrees of preterm births is important as decreasing length of gestation is associated with increased risk of mortality, disability, and morbidity across the lifespan (82). Also, considering preterm births as not being homogeneous in causes and consequences was highlighted by others (83, 84) as an important approach when interpreting such results, but a major limitation in current evidence (6, 7, 34).

Our main analysis suggested that very preterm have an increased odds of overweight in adolescence compared with full term peers. Despite heterogeneity in characteristics for the cohorts (ALSPAC, DNBC, NFBC1986 and the Raine Study) included for this age-band, our supplementary analyses addressed robustness in our findings. Our results are further supported by findings from two comparable studies conducted in Finland and Australia, where an increased odds of overweight was reported in preterm aged 23 and 35 years, respectively (81, 85).

In summary, this study sheds new light on factors influencing BMI and the odds of developing overweight from infancy through adolescence. Our analysis revealed that although preterm infants are relatively small at birth, they reach similar levels of BMI and odds of overweight by adolescence as full term counterparts. The underlying mechanisms from the current observational data are unknown. However, in accordance with previous findings, our pattern of results suggests that preterm infants may be at an increased odds of overweight later in life, even though mean BMI in preterm and full term are similar. In addition, it should be noted that mediating exposures such as birth weight, congenital anomalies and breast feeding practices may also affect the relationship between GA and later body size.

### Strengths and limitations

An important strength in the current study is the large sample size with information on more than 250,000 mother-child dyads, from 16 prospective pregnancy and birth cohorts in Europe, North America, and Australasia. We used comprehensive obstetric and maternal data as well as multiple BMI measurements following birth through adolescence, which allowed us to adjust analyses. Additionally, the large sample size enabled us to assess associations successively using clinical categories of preterm birth to age 19 years. We also examined the robustness of our findings performing several sensitivity analyses. Furthermore, the federated analysis approach using DataSHIELD proves a key advantage since it enables identical and reproducible analysis across multiple cohorts (39, 86, 87).

The limitations include the considerable variations that exist in the measurement and availability of both exposures, covariates, and outcomes. However, this was explored by meta-regressions on multiple covariates showing that study characteristics were independently associated with between-study heterogeneity only in the associations of GA with BMI. Age at measurement was the main contributor to heterogeneity in early infancy, but not in childhood and adolescence. This suggests that GA is important for BMI in early life, but attenuates consistently as children get older. In late childhood and adolescence, maternal education and maternal smoking in pregnancy were independently associated with the observed heterogeneity.

Residual confounding may be another limitation in this study as the confounders are harmonised across studies, which gives the lowest common denominator. Several large cohorts (DNBC, MOBA, NFBC1986, the Raine study) had no available information on maternal ethnic background (Table S1), which could bias our results. However, we had reports that the cohorts were homogeneous (>95% western) (45, 52, 53, 56), hence we do not assume this affected our findings.

As survival rates and postnatal treatment for preterm infants have improved in the last 20 years (3), distribution and characteristics of GA in the earliest cohorts are likely to differ from that in populations born more recently, with the former potentially being more selected and healthy later (34, 88). We do, however, not assume this would affect our overall estimates markedly.

## Conclusions

Higher GA is potentially clinically important for higher BMI in infancy, while the association attenuates consistently with age. By adolescence, preterm children have on average a similar mean BMI to those born full term.

## Data Availability

Data Availability Statement The data used for this study is third-party data, and without legally permission of distribution or public sharing. Information about data access and governance for this study is explained in detail in a peer-reviewed scientific paper by Prof. Vincent Jaddoe et al (2020): “The LifeCycle Project?EU Child Cohort Network: a federated analysis infrastructure and harmonized data of more than 250,000 children and parents”. The principal investigators or home institutions administer permission to the data for external researchers: hence, access to the data is conditional on reasonable request and with approval by each cohort. A description of the data set and third-party sources are listed in Supplementary Materials, and displayed online at the EU Child Cohort Variable Catalogue (https://data-catalogue.molgeniscloud.org/catalogue/catalogue/#/) and the Maelstrom Catalogue (https://www.maelstrom-research.org/page/catalogue).

## Acknowledgement

The authors would like to acknowledge everyone in LifeCycle and EUCAN-Connect who have supported and contributed to each cohort included in the study. In addition, acknowledgements are sent to the DataSHIELD team. Please see S2 Appendix for list of cohort-specific acknowledgements.

## Author Contribution

**Conceptualization:** Johan L. Vinther, Anne-Marie Nybo Andersen, Claus T. Ekstrøm, Thorkild I.A. Sørensen, Deborah Lawlor

**Data Curation:** Johan L. Vinther, Tim Cadman, Demetris Avraam, Thorkild I.A. Sørensen, Claus T. Ekstrøm, Ahmed Elhakeem, Ana C. Santos, Angela Pinot de Moira, Barbara Heude, Carmen Iñiguez, Costanza Pizzi, Elinor Simons, Ellis Voerman, Eva Corpeleijn, Faryal Zariouh, Gilian Santorelli, Hazel M. Inskip, Henrique Barros, Jennie Carson, Jennifer R. Harris, Johanna L Nader, Justiina Ronkainen, Loreto Santa-Marina, Lucinda Calas, Luise Cederkvist, Maja Popovic, Marie-Aline Charles, Martine Vrijheid, Meghan Azad, Padmaja Subbarao, Paul Burton, Puishkumar J. Mandhane, Rae-Chi Huang, Rebecca C. Wilson, Sido Haakma, Silvia Fernandez, Stuart Turvey, Susana Santos, Suzanne C. Tough, Sylvain Sebert, Tanis Fenton, Theo Moraes, Theodosia Salika, Vincent W.V. Jaddoe, Deborah A. Lawlor, Anne-Marie Nybo Andersen

**Formal Analysis:** Johan L. Vinther, Tim Cadman, Demetris Avraam

**Funding Acquisition:** Vincent Jaddoe, Martine Vrijheid, Marie-Aline Charles, Jennifer R. Harris, Deborah A. Lawlor, Anne-Marie Nybo Andersen, Hazel Inskip, Rae-Chi Huang

**Methodology:** Johan L. Vinther, Anne-Marie Nybo Andersen, Tim Cadman, Demetris Avraam, Claus T. Ekstrøm, Deborah A. Lawlor, Thorkild I.A. Sørensen

**Project Administration:** Johan L. Vinther

**Software:** Johan L. Vinther, Tim Cadman, Demetris Avraam, Sido Haakma, Paul Burton

**Supervision:** Anne-Marie Nybo Andersen, Deborah A. Lawlor, Claus T. Ekstrøm

**Visualization:** Johan L. Vinther

**Writing – Original Draft Preparation:** Johan L. Vinther, Anne-Marie Nybo Andersen, Tim Cadman, Demetris Avraam, Claus T. Ekstrøm, Thorkild I.A. Sørensen, Deborah A. Lawlor

**Writing – Review & Editing:** Johan L. Vinther, Tim Cadman, Demetris Avraam, Thorkild I.A. Sørensen, Claus T. Ekstrøm, Ahmed Elhakeem, Ana C. Santos, Angela Pinot de Moira, Barbara Heude, Carmen Iñiguez, Costanza Pizzi, Elinor Simons, Ellis Voerman, Eva Corpeleijn, Faryal Zariouh, Gilian Santorelli, Hazel M. Inskip, Henrique Barros, Jennie Carson, Jennifer R. Harris, Johanna L Nader, Justiina Ronkainen, Katrine Strandberg-Larsen, Loreto Santa-Marina, Lucinda Calas, Luise Cederkvist, Maja Popovic, Marie-Aline Charles, Martine Vrijheid, Meghan Azad, Padmaja Subbarao, Paul Burton, Puishkumar J. Mandhane, Rae-Chi Huang, Rebecca C. Wilson, Sido Haakma, Sílvia Fernández-Barrés, Stuart Turvey, Susana Santos, Suzanne C. Tough, Sylvain Sebert, Tanis Fenton, Theo Moraes, Theodosia Salika, Vincent W.V. Jaddoe, Deborah A. Lawlor, Anne-Marie Nybo Andersen

## SUPPORTING INFORMATION

**S1 Table.**
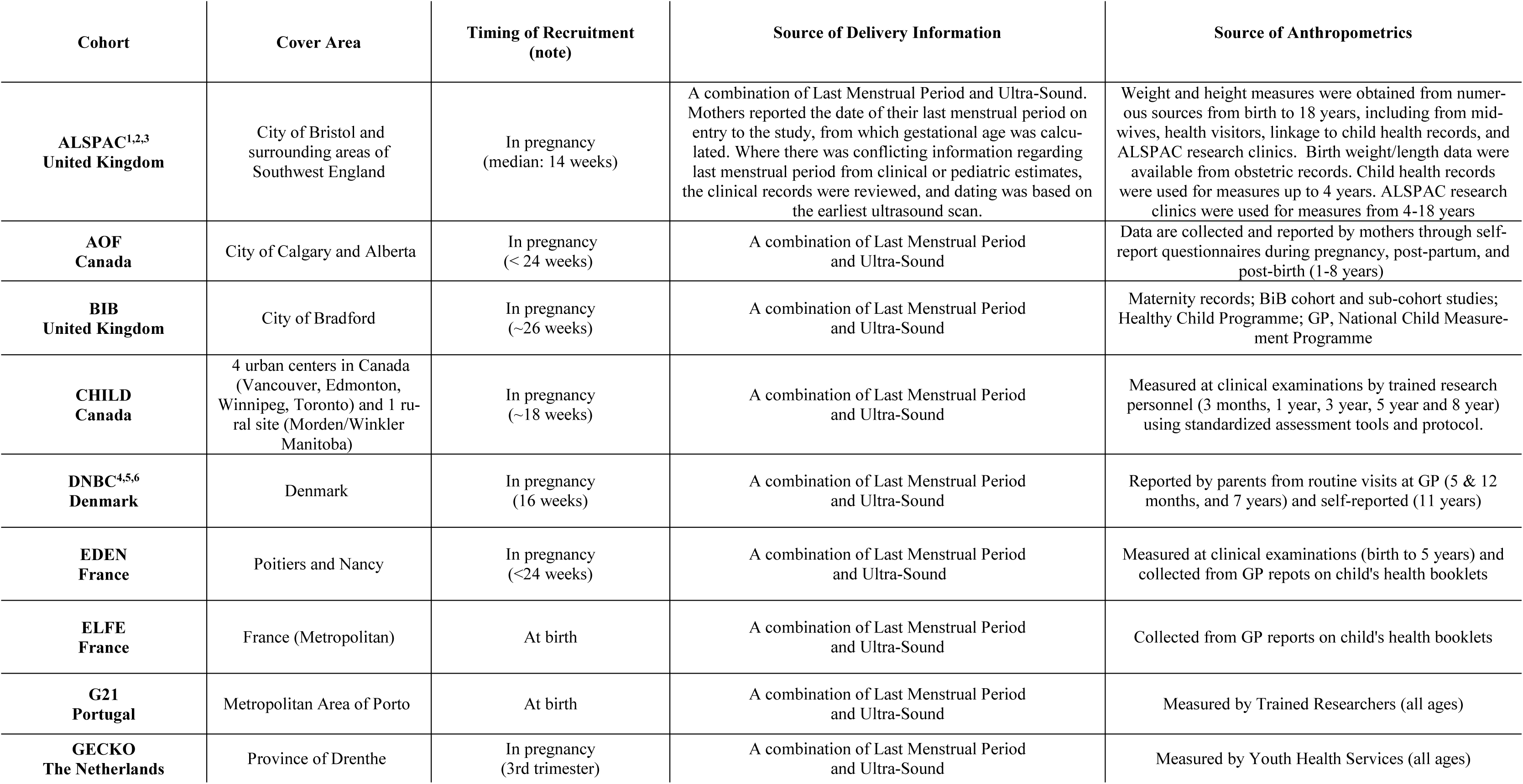
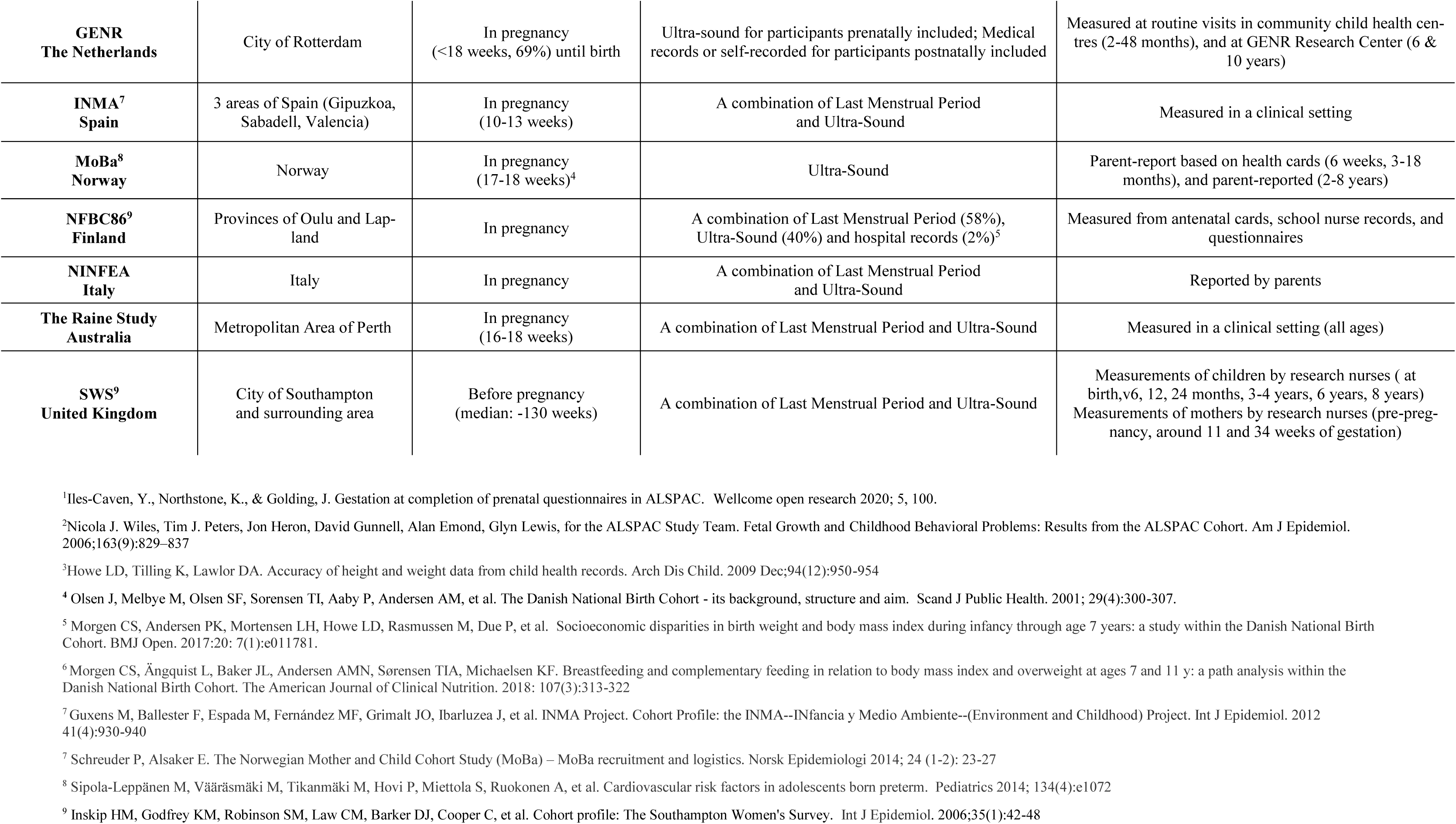
Cohort-specific study characteristics and information on exposure and outcome measurements.

**S2 Table.**
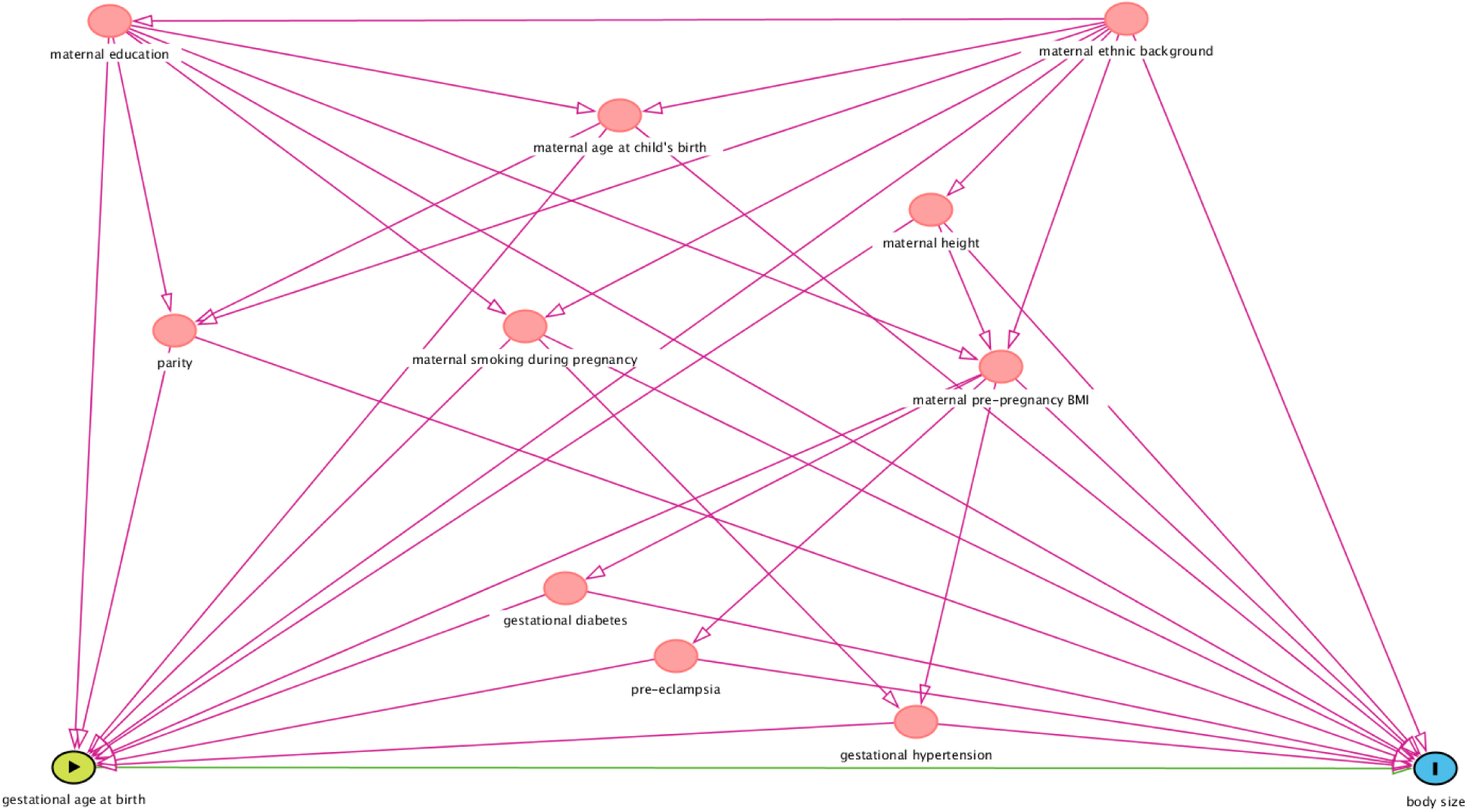
Directed Acyclic Graph for the association between GA and Body Size.

**S3 Table.** Percentage of missing values for cohort-specific baseline characteristics.

|  | N | Maternal Age at Child's Birth | Maternal Education | Maternal Ethnicity | Maternal Height | Pre-pregnancy BMI | Smoking in Pregnancy | Gestational Diabetes | Gestational Hypertension | Maternal Preeclampsia | Parity |
| --- | --- | --- | --- | --- | --- | --- | --- | --- | --- | --- | --- |
| ALSPAC<br>United Kingdom | 10,542 | 8.8 | 5.3 | 6.2 | 7.0 | 15.4 | 11.5 | 5.7 | 2.4 | 0.4 | 4.3 |
| AOF<br>Canada | 2,263 | 0.0 | 0.2 | 0.4 | 0.5 | 1.3 | 0.0 | 0.0 | 0.0 | 0.0 | 0.8 |
| BIB<br>United Kingdom | 13,097 | 0.0 | 23.8 | 17.2 | 18.9 | 64.5 | 17.3 | 6.6 | 18.3 | 18.3 | 3.7 |
| CHILD<br>Canada | 2,984 | 0.0 | 3.4 | 0.9 | 3.1 | 31.8 | 2.3 | 0.0 | 16.3 | 2.3 | 0.1 |
| DNBC<br>Denmark | 81,117 | 0.0 | 11.1 |  | 5.0 | 6.4 | 1.5 | 0.0 | 25.3 | 1.6 | 0.0 |
| EDEN<br>France | 1,765 | 0.0 | 0.6 | 13.8 | 1.4 | 2.0 | 0.3 | 0.1 | 0.1 | 1.1 | 0.2 |
| ELFE<br>France | 15,506 | 0.3 | 0.0 | 6.7 | 0.3 | 1.3 | 0.9 | 3.9 | 1.8 | 1.8 | 1.2 |
| G21<br>Portugal | 6,439 | 0.1 | 0.6 | 5.2 | 0.2 | 2.0 | 43.8 | 0.5 | 0.5 | 0.0 | 1.8 |
| GECKO<br>The Netherlands | 2,768 | 0.9 | 7.0 | 6.8 | 5.3 | 8.5 | 1.0 | 13.8 | 2.6 | 8.2 | 1.5 |
| GEN-R<br>The Netherlands | 8,641 | 0.0 | 10.3 | 5.4 | 3.7 | 23.4 | 14.0 | 3.9 | 12.1 | 12.1 | 2.8 |
| INMA<br>Spain | 1,936 | 0.0 | 2.0 | 1.8 | 0.9 | 0.9 | 1.2 | 7.1 |  |  | 4.5 |
| MoBa<br>Norway | 86,553 | 0.2 | 5.9 |  | 2.2 | 3.5 | 0.0 | 1.1 | 4.1 | 4.1 | 1.1 |
| NFBC1986<br>Finland | 8,325 | 0.0 | 12.3 |  | 0.8 | 2.2 | 0.5 |  | 19.1 | 20.3 | 0.3 |
| NINFEA<br>Italy | 6,514 | 0.0 | 0.7 |  | 2.1 | 2.2 | 1.2 | 8.8 | 8.4 | 8.2 | 4.8 |
| The Raine study<br>Australia | 2,443 | 0.2 | 0.5 | 0.0 | 0.0 | 2.8 | 0.0 | 0.0 | 2.7 | 2.7 | 0.0 |
| SWS<br>United Kingdom | 3,007 | 0.0 | 0.3 | 0.3 | 0.5 | 0.9 | 4.6 | 0.0 | 0.0 | 0.0 | 0.1 |

**Note:** Empty cells represent no available data
**Abbreviations:** N = Sample Size, BMI = Body Mass Index

**S4 Table.** Results of individual variable meta-regression models showing values of β, se(β), and the significance of β for each study characteristic.

| **Age-band** | **Study Characteristics** | **$\beta$** | **$se(\beta)$** | **p-value** |
| --- | --- | --- | --- | --- |
| **Early infancy (>0.0-0.5 years)** | *Age at measurement, months* | **-0.0296** | 0.0076 | <0.01 |
|  | *Sex, female (%)* | **0.0016** | 0.0096 | 0.87 |
|  | *Maternal education, low (%)* | **0.0011** | 0.0006 | 0.10 |
|  | *Maternal smoking in pregnancy, yes (%)* | **-0.0005** | 0.0011 | 0.68 |
| **Mid-childhood (>5.0-9.0 years)** | *Age at measurement, months* | **0.0006** | 0.0005 | 0.27 |
|  | *Sex, female (%)* | **0.0042** | 0.0047 | 0.38 |
|  | *Maternal education, low (%)* | **0.0002** | 0.0003 | 0.59 |
|  | *Maternal smoking in pregnancy, yes (%)* | **0.0000** | 0.0005 | 0.94 |
| **Late childhood (>9.0-14.0 years)** | *Age at measurement, months* | **0.0001** | 0.0005 | 0.83 |
|  | *Sex, female (%)* | **0.0019** | 0.0071 | 0.77 |
|  | *Maternal education, low (%)* | **0.0009** | 0.0005 | 0.05 |
|  | *Maternal smoking in pregnancy, yes (%)* | **0.0002** | 0.0007 | 0.74 |
| **Adolescence (>14.0-19.0 years)** | *Age at measurement, months* | **-0.0007** | 0.0012 | 0.58 |
|  | *Sex, female (%)* | **-0.0020** | 0.0034 | 0.55 |
|  | *Maternal education, low (%)* | **0.0012** | 0.0005 | <0.01 |
|  | *Maternal smoking in pregnancy, yes (%)* | **0.0074** | 0.0030 | 0.01 |

**S1 Fig.**
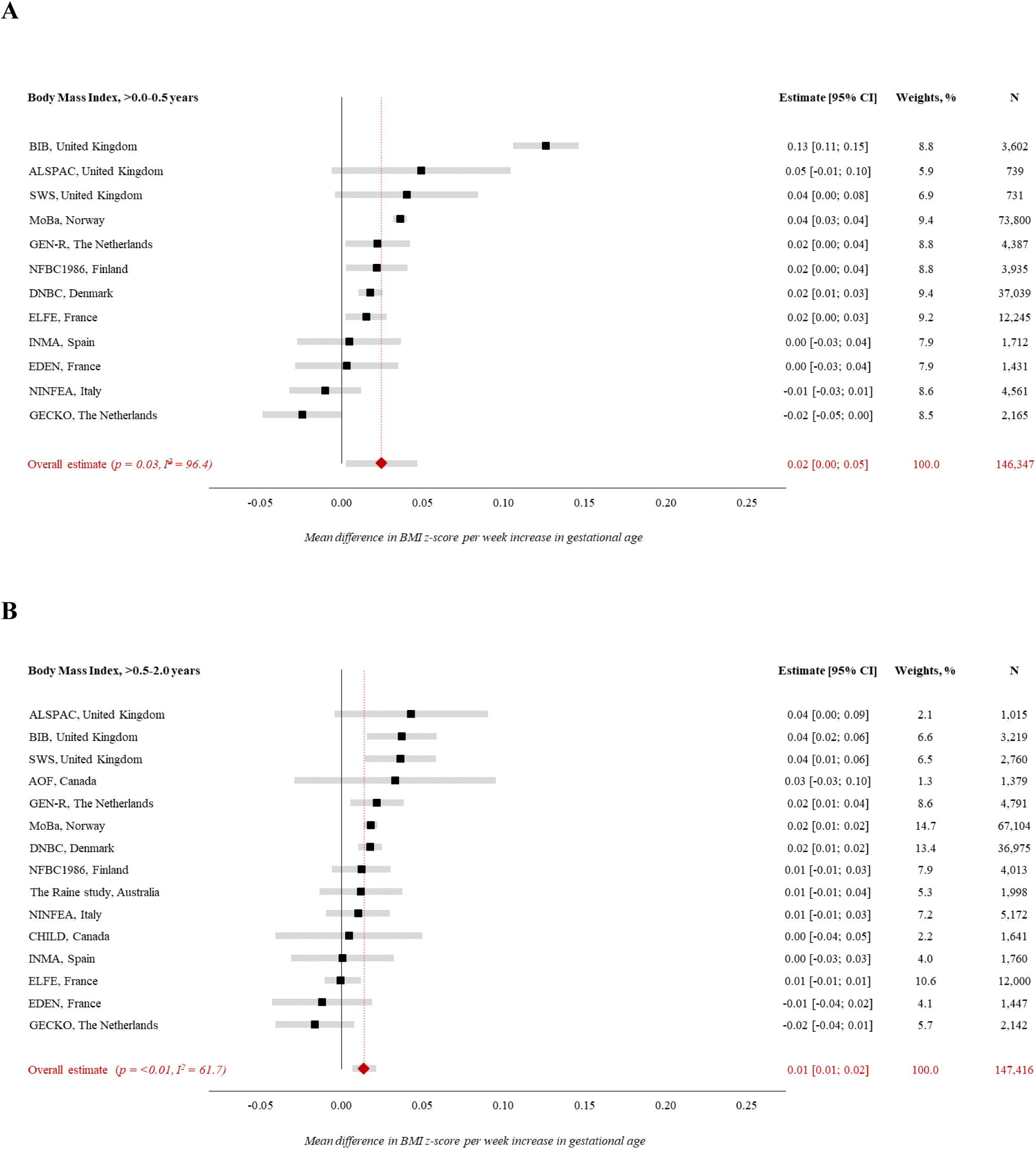
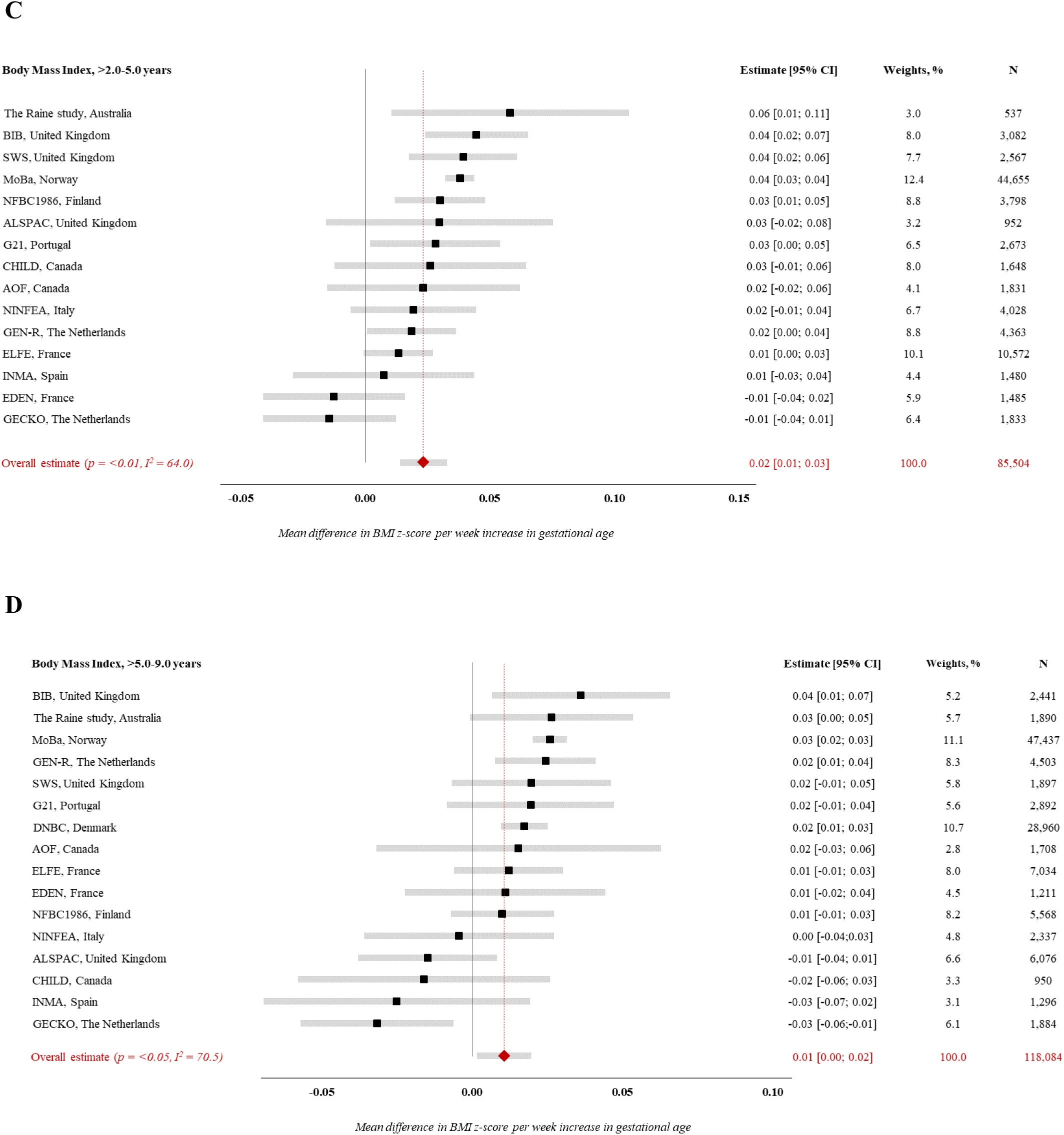
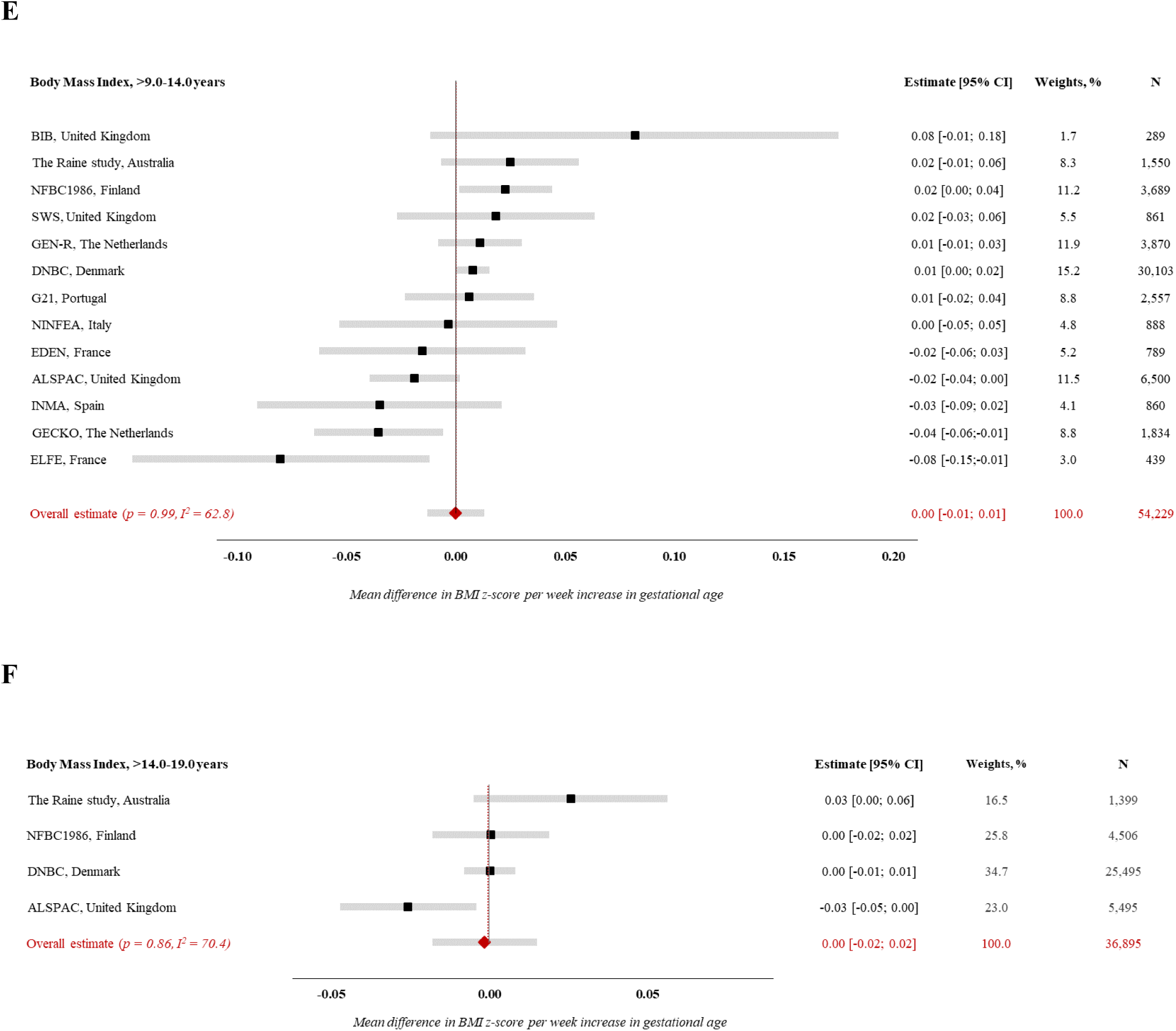
Forest plot of associations between GA (completed weeks) and BMI z-score. Estimates from study-specific linear regression models were assigned weights under the random-effects model to attain overall estimates. Results reflect mean differences in BMI z-score per week increase in gestational age at birth in (A) early infancy (>0.0-0.5 years), (B) late infancy (>0.5-2.0 years), (C) early childhood (>2.0-5.0 years), (D) mid-childhood (>5.0-9.0 years), (E) late childhood (>9.0-14.0 years), (F) adolescence (>14.0-19.0 years) Models are adjusted for sex of child, maternal age at child’s birth, maternal education, maternal height, maternal pre-pregnancy BMI, maternal smoking during pregnancy, parity, maternal ethnic background, gestational diabetes, gestational hypertension, pre-eclampsia. Specific variables were not available in few cohorts (see Table 1), hence not adjusted for.

**S2 Fig.**
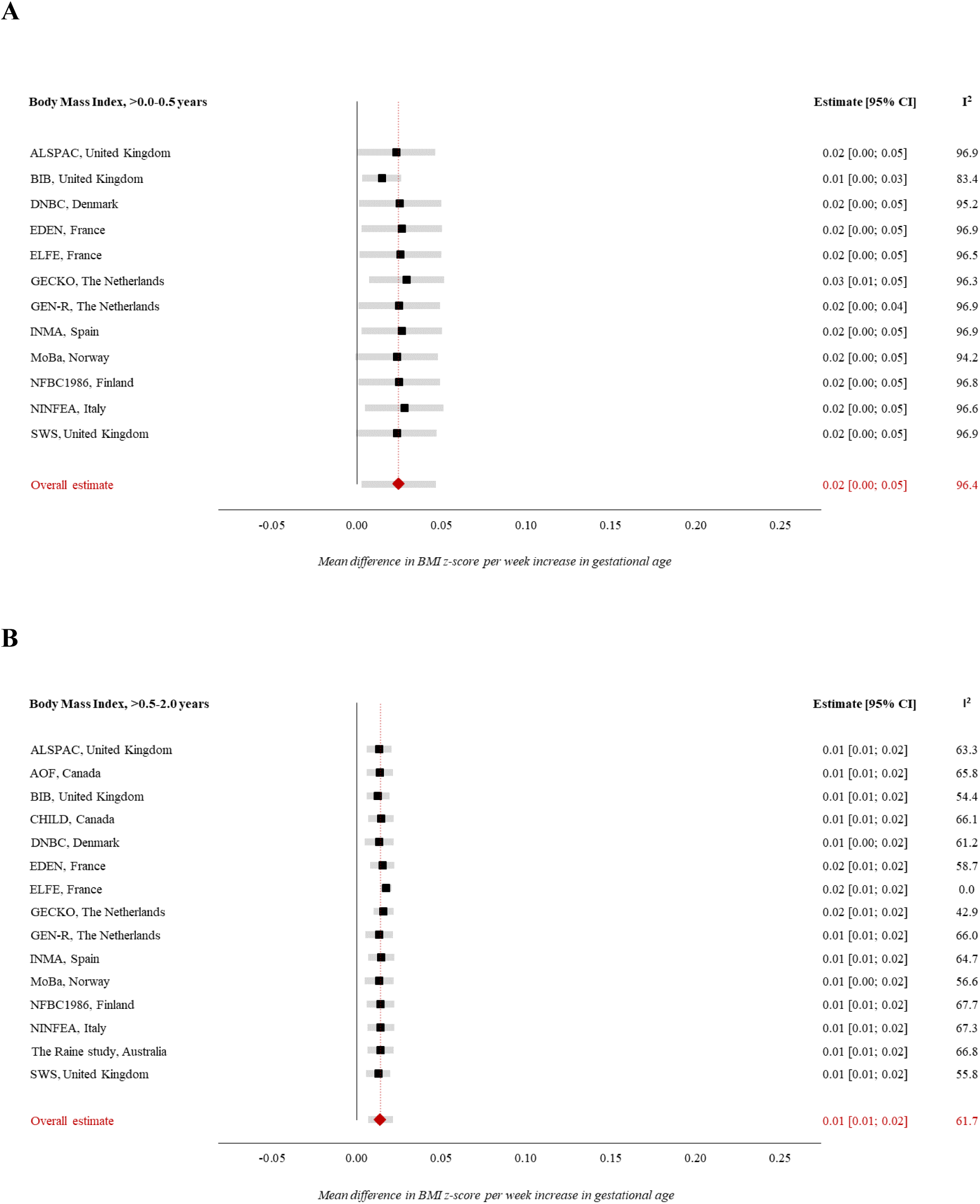
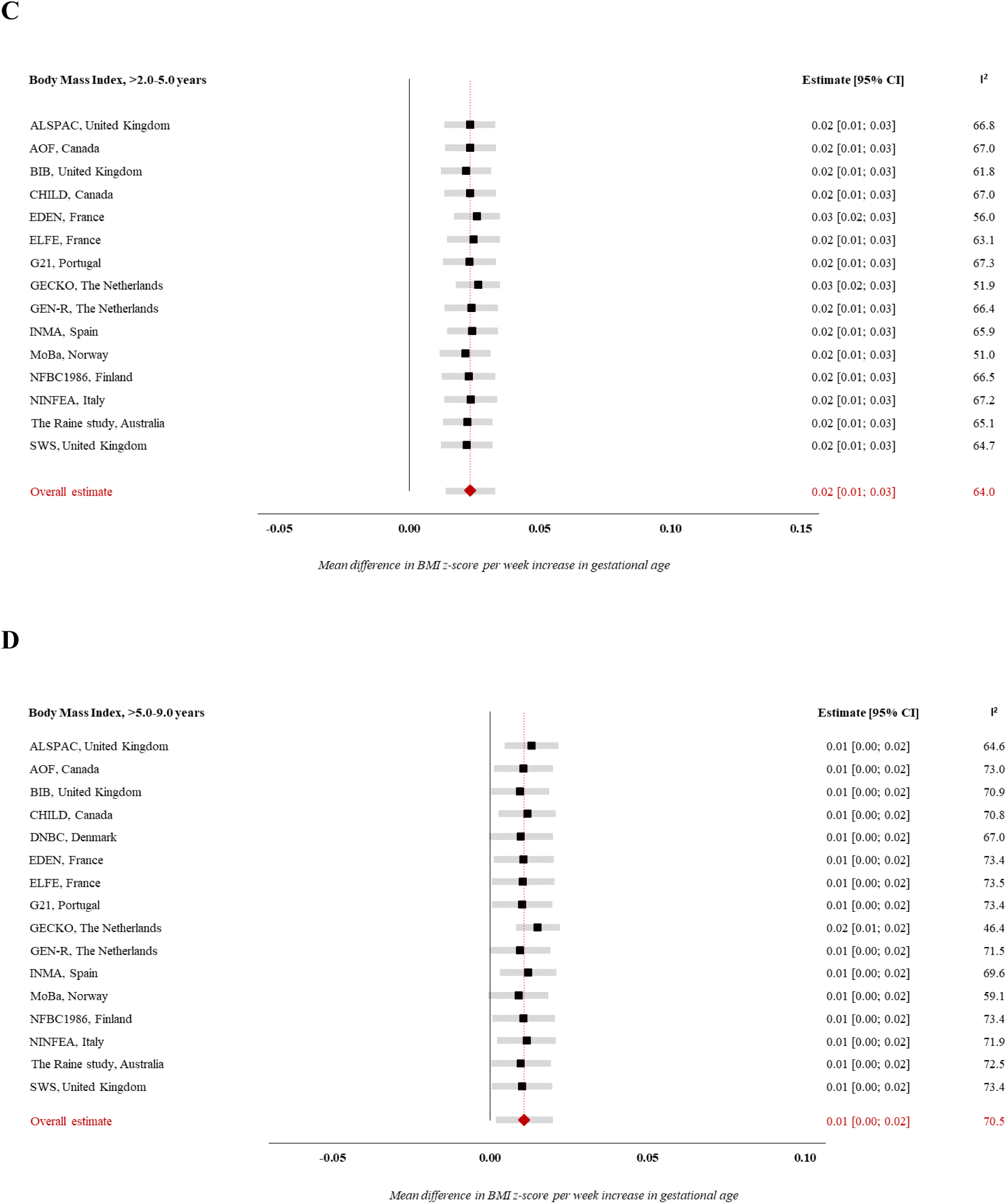
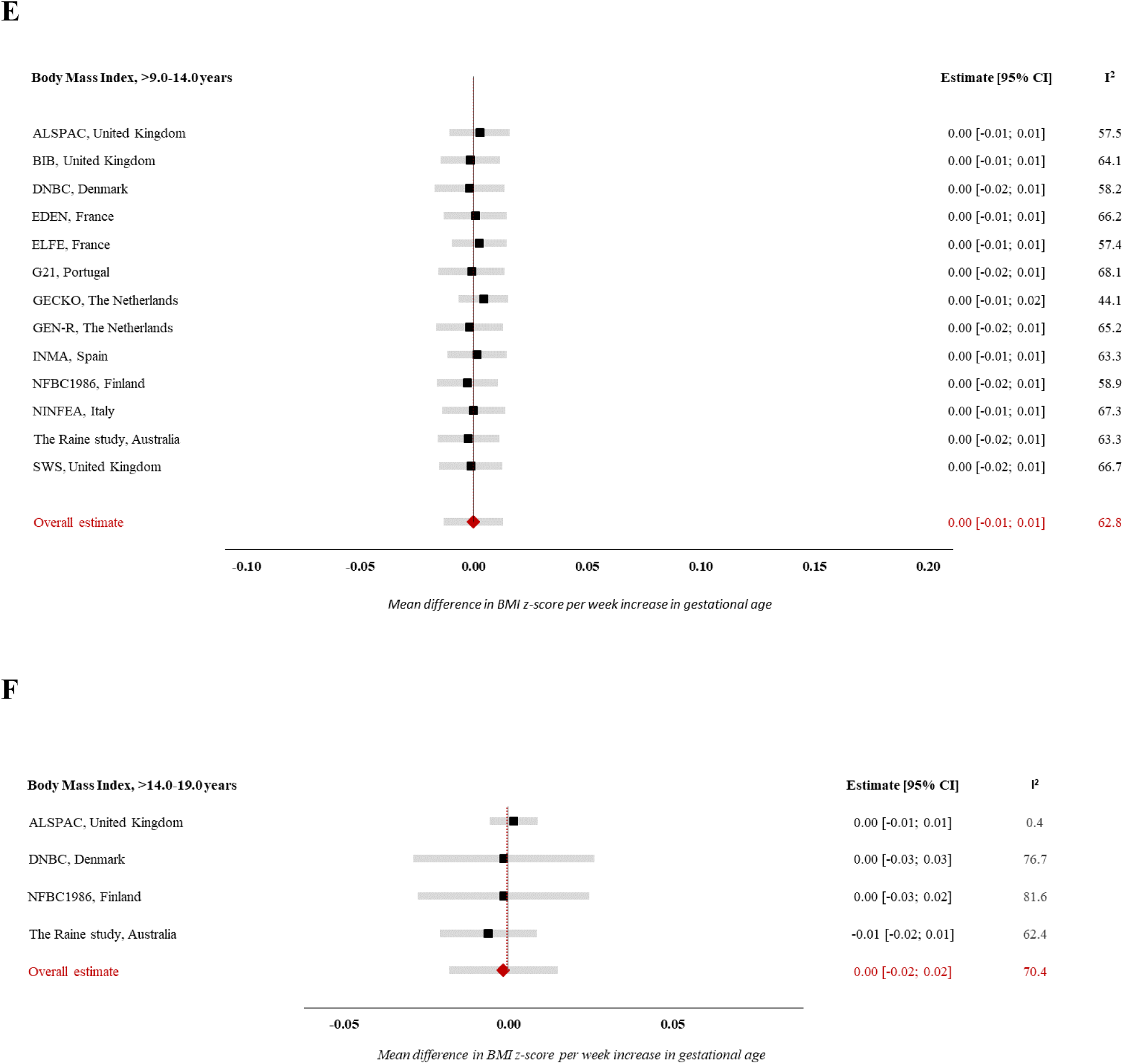
Forest plot of ‘Leave-one-out’ analysis for the association between GA (in weeks) and BMI z-score. Estimates from study-specific linear regression models were assigned weights under the random-effects model to attain overall estimates. Results reflect mean differences in BMI z-score per week increase in gestational age at birth in (A) early infancy (>0.0-0.5 years), (B) late infancy (>0.5-2.0 years), (C) early childhood (>2.0-5.0 years), (D) mid-childhood (>5.0-9.0 years), (E) late childhood (>9.0-14.0 years), (F) adolescence (>14.0-19.0 years) Models are adjusted for sex of child, maternal age at child’s birth, maternal education, maternal height, maternal pre-pregnancy BMI, maternal smoking during pregnancy, parity, maternal ethnic background, gestational diabetes, gestational hypertension, pre-eclampsia. Specific variables were not available in few cohorts (see Table 1), hence not adjusted for.

**S3 Fig.**
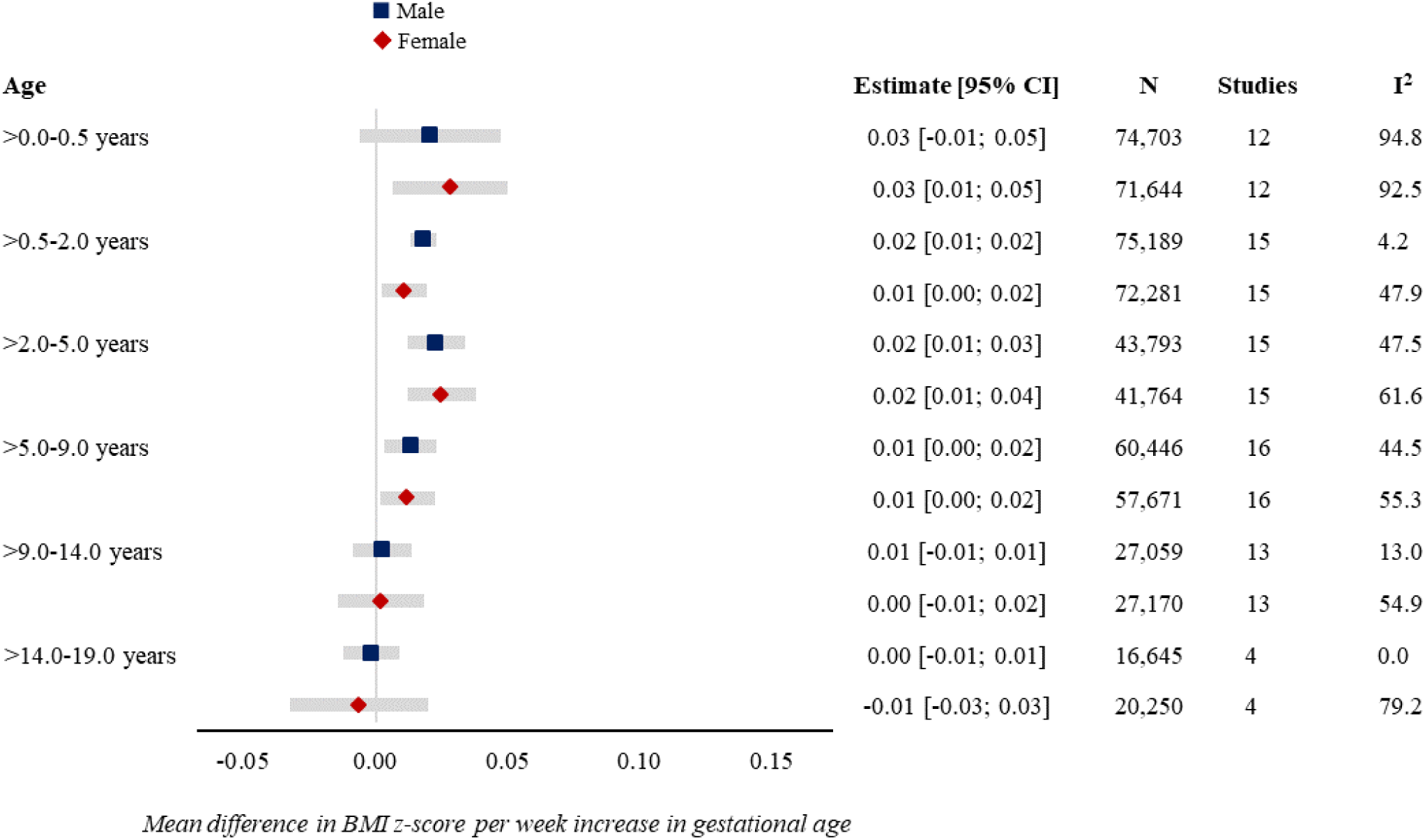
Forest plot of associations between GA (completed weeks) and BMI z-score by sex. Estimates from study-specific linear regression models were assigned weights under the random-effects model to attain overall estimates. Results reflect mean differences in BMI z-score per week increase in gestational age at birth in early infancy (>0.0-0.5 years), late infancy (>0.5-2.0 years), early childhood (>2.0-5.0 years), mid-childhood (>5.0-9.0 years), late childhood (>9.0-14.0 years), adolescence (>14.0-19.0 years). Models are adjusted for sex of child, maternal age at child’s birth, maternal education, maternal height, maternal pre-pregnancy BMI, maternal smoking during pregnancy, parity, maternal ethnic background, gestational diabetes, gestational hypertension, pre-eclampsia. Specific variables were not available in few cohorts (see Table 1), hence not adjusted for.

**S4 Fig.**
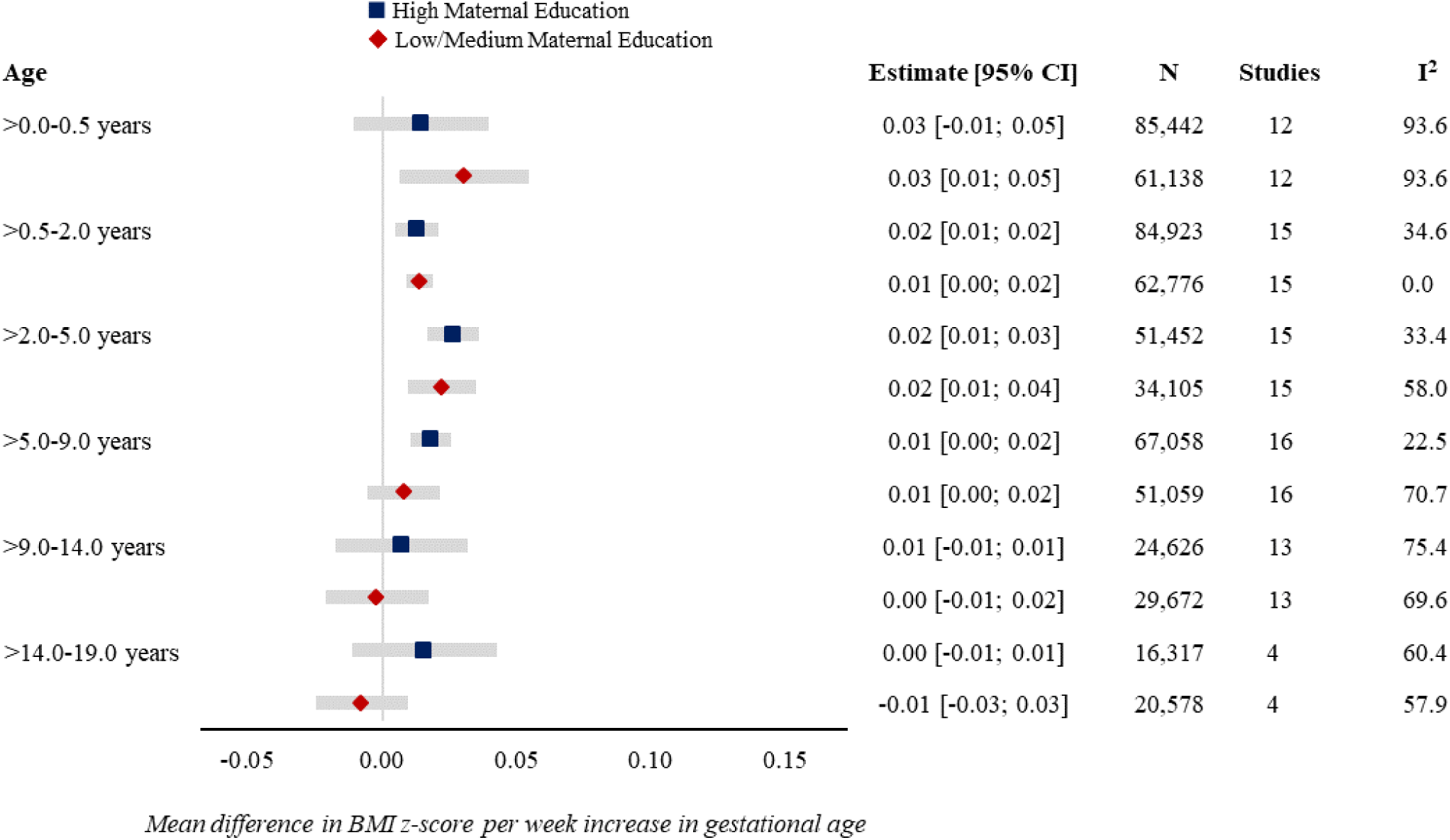
Forest plot of associations between GA (completed weeks) and BMI z-score by maternal education. Estimates from study-specific linear regression models were assigned weights under the random-effects model to attain overall estimates. Results reflect mean differences in BMI z-score per week increase in gestational age at birth in early infancy (>0.0-0.5 years), late infancy (>0.5-2.0 years), early childhood (>2.0-5.0 years), mid-childhood (>5.0-9.0 years), late childhood (>9.0-14.0 years), adolescence (>14.0-19.0 years). Models are adjusted for sex of child, maternal age at child’s birth, maternal education, maternal height, maternal pre-pregnancy BMI, maternal smoking during pregnancy, parity, maternal ethnic background, gestational diabetes, gestational hypertension, pre-eclampsia. Specific variables were not available in few cohorts (see Table 1), hence not adjusted for.

**S5 Fig.**
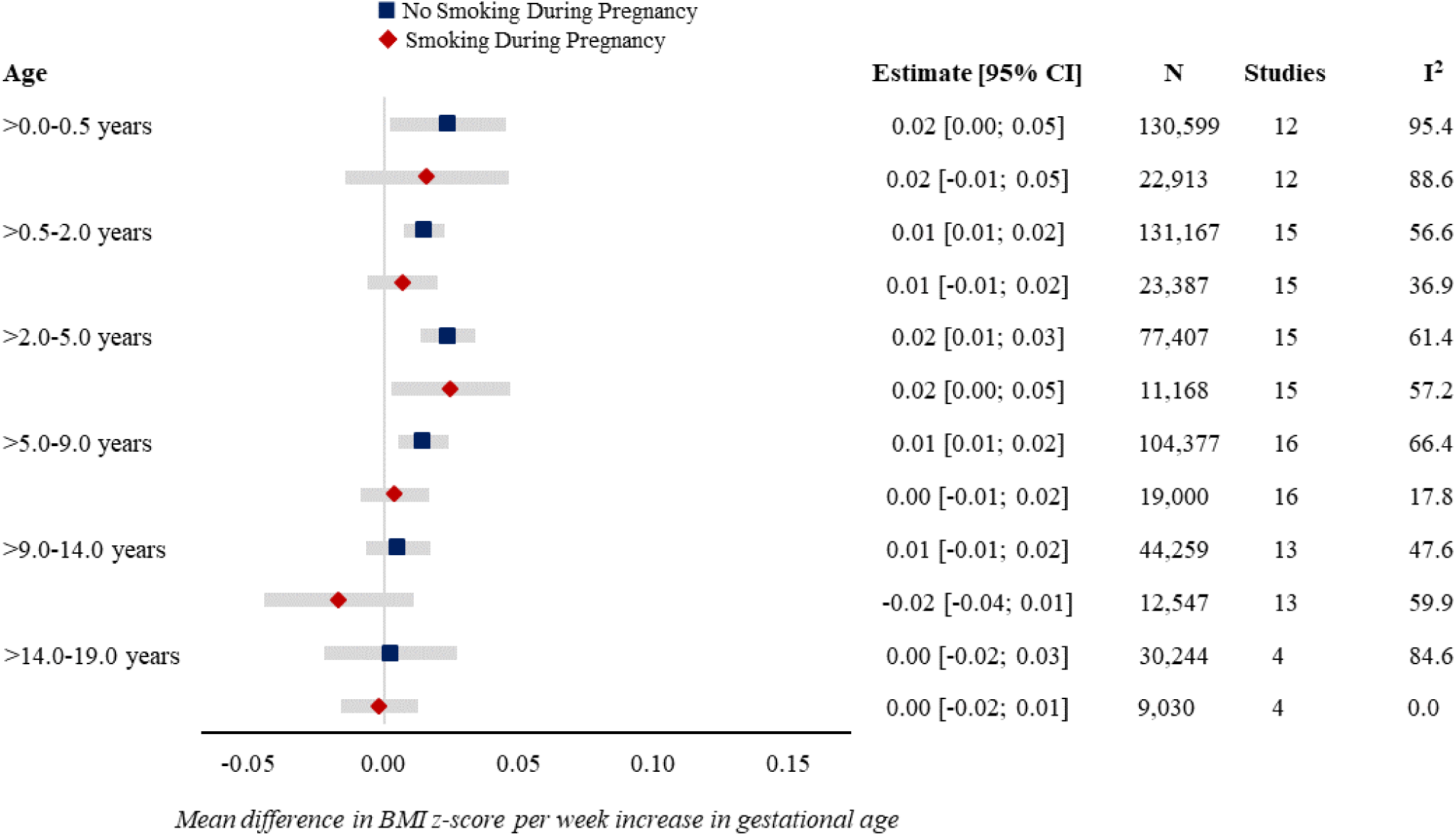
Forest plot of associations between GA (completed weeks) and BMI z-score by maternal smoking in pregnancy. Estimates from study-specific linear regression models were assigned weights under the random-effects model to attain overall estimates. Results reflect mean differences in BMI z-score per week increase in gestational age at birth in early infancy (>0.0-0.5 years), late infancy (>0.5-2.0 years), early childhood (>2.0-5.0 years), mid-childhood (>5.0-9.0 years), late childhood (>9.0-14.0 years), adolescence (>14.0-19.0 years). Models are adjusted for sex of child, maternal age at child’s birth, maternal education, maternal height, maternal pre-pregnancy BMI, maternal smoking during pregnancy, parity, maternal ethnic background, gestational diabetes, gestational hypertension, pre-eclampsia. Specific variables were not available in few cohorts (see Table 1), hence not adjusted for.

**S6 Fig.**
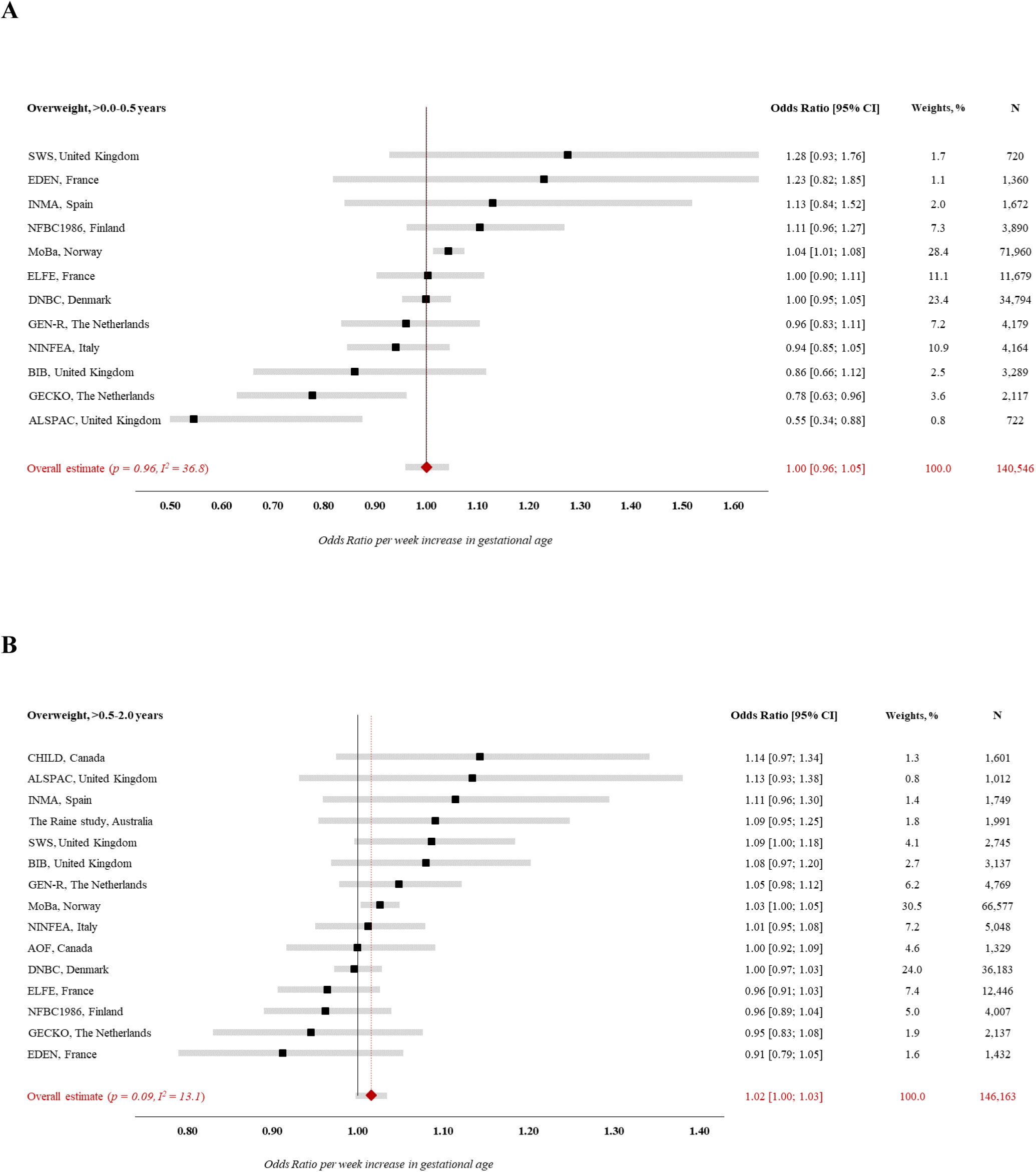
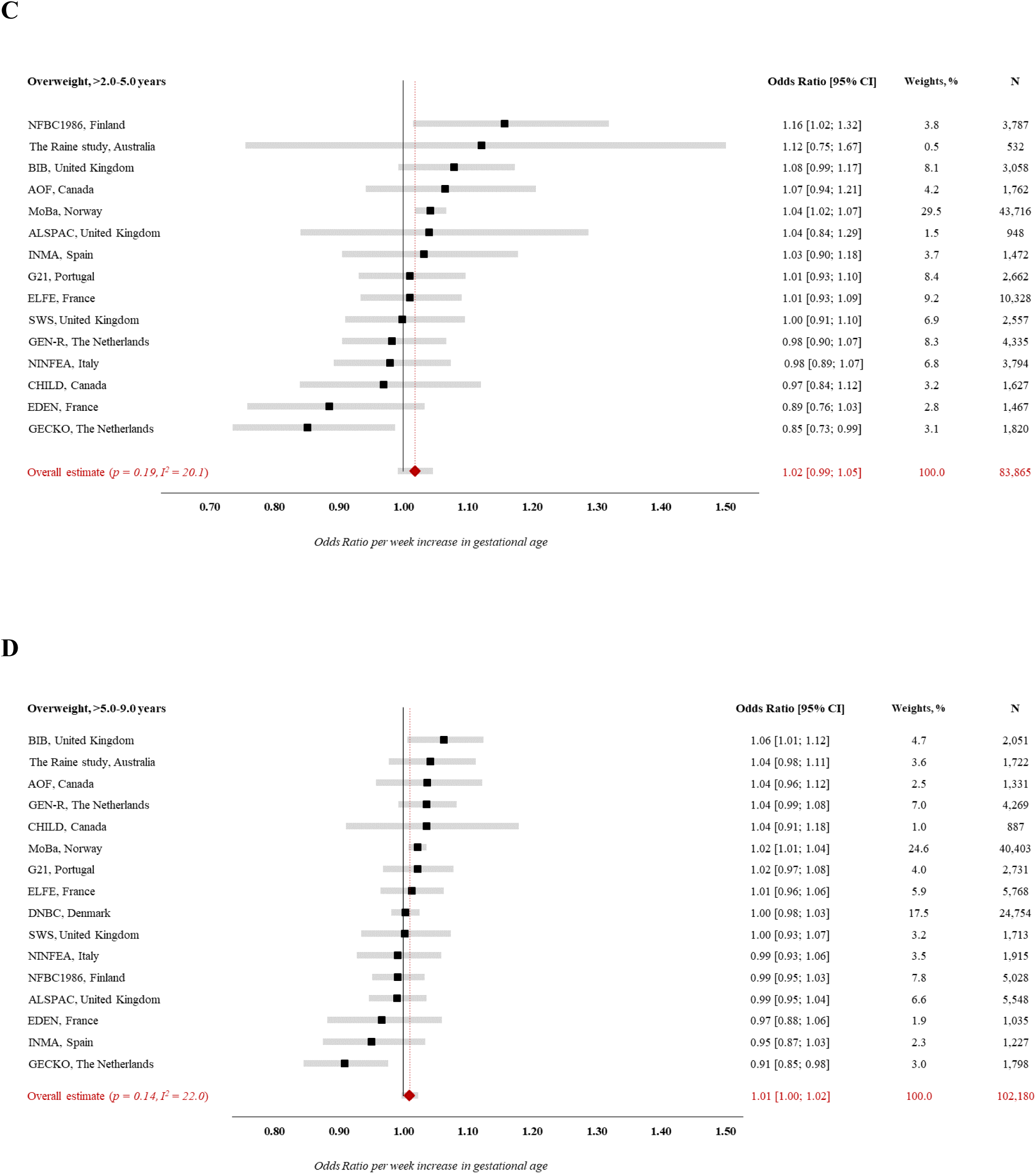
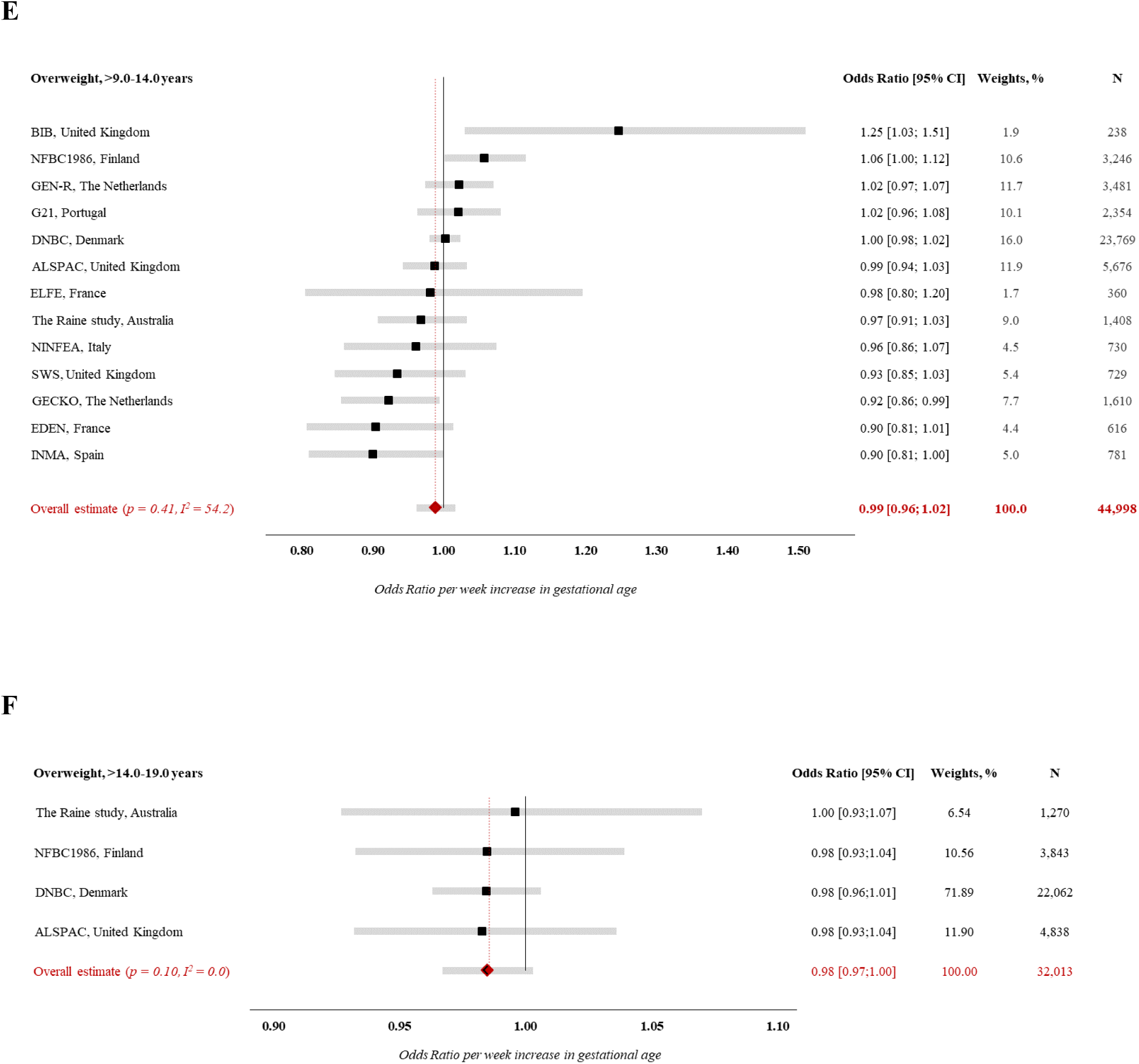
Forest plot of associations between GA (completed weeks) and odds of overweight. Estimates from study-specific logistic regression models were assigned weights under the random-effects model to attain overall estimates. Results reflect odds ratio of overweight (vs. normal weight) per week increase in gestational age at birth in (A) early infancy (>0.0-0.5 years), (B) late infancy (>0.5-2.0 years), (C) early childhood (>2.0-5.0 years), (D) mid-childhood (>5.0-9.0 years), (E) late childhood (>9.0-14.0 years), (F) adolescence (>14.0-19.0 years) Models are adjusted for sex of child, maternal age at child’s birth, maternal education, maternal height, maternal pre-pregnancy BMI, maternal smoking during pregnancy, parity, maternal ethnic background, gestational diabetes, gestational hypertension, pre-eclampsia. Specific variables were not available in few cohorts (see Table 1), hence not adjusted for.

**S7 Fig.**
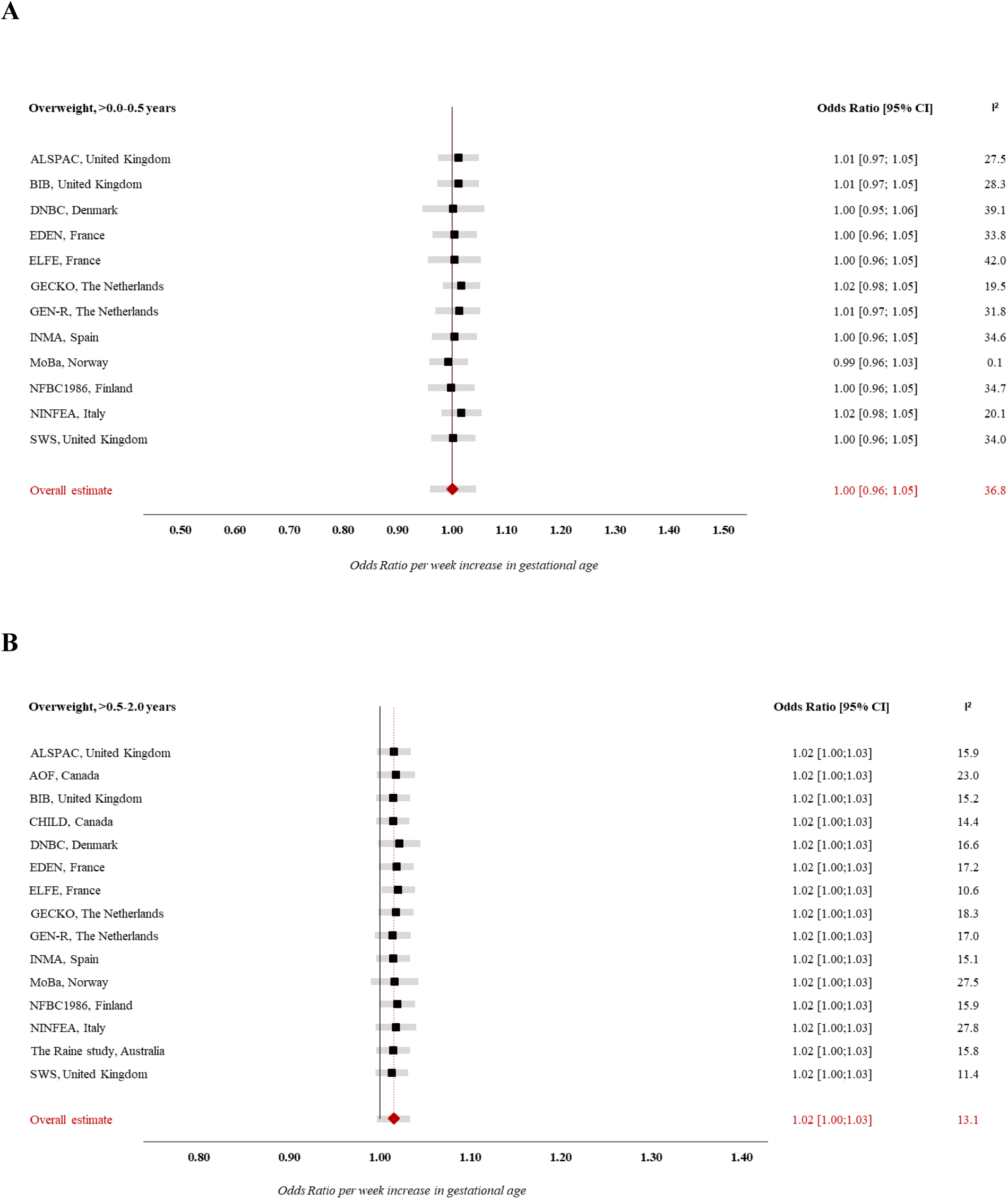
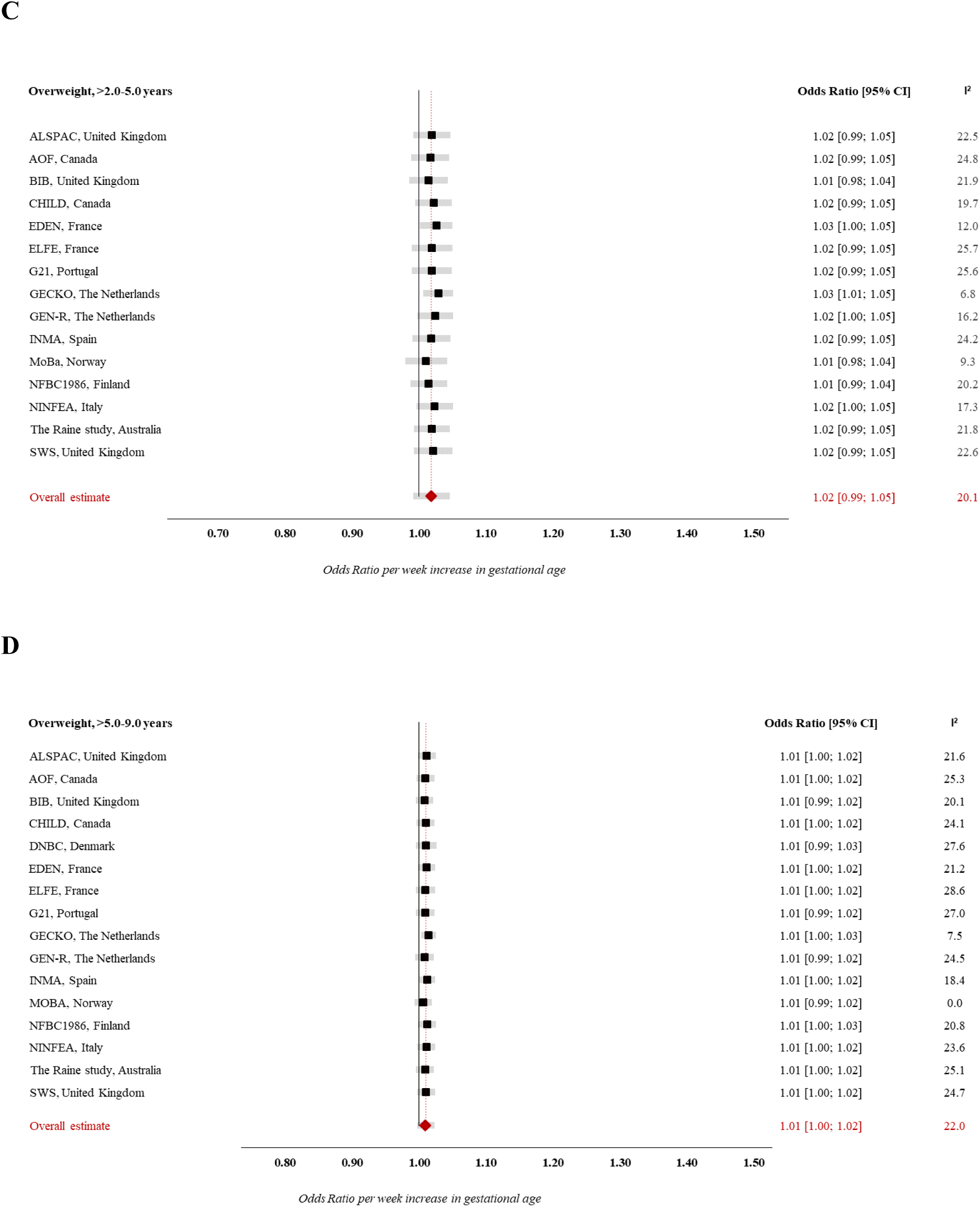
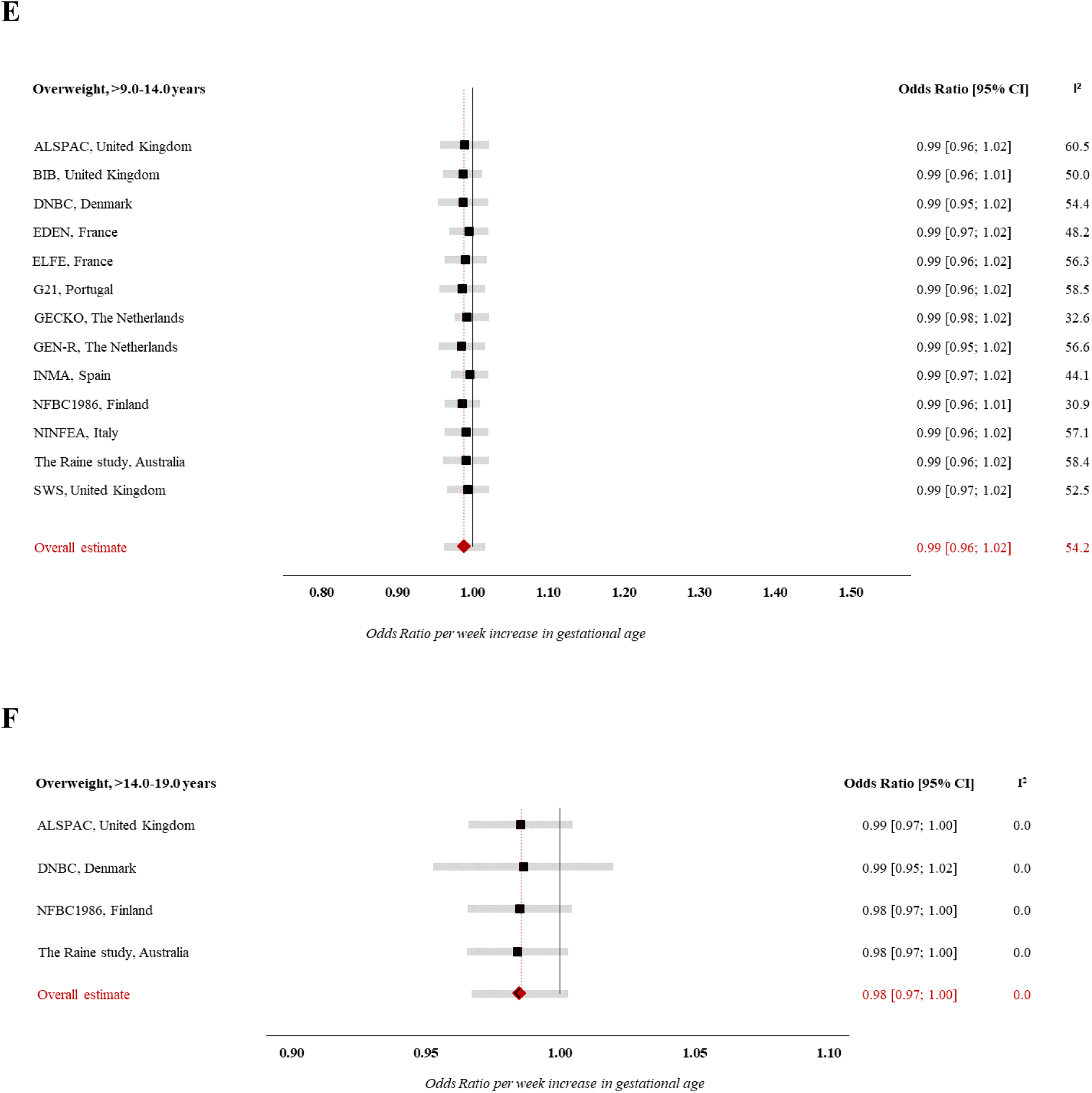
Forest plot of ‘Leave-one-out’ analysis for the association between GA (completed weeks) and odds of overweight. Estimates from study-specific logistic regression models were assigned weights under the random-effects model to attain overall estimates. Results reflect odds ratio of overweight (vs. normal weight) per week increase in gestational age at birth in (A) early infancy (>0.0-0.5 years), (B) late infancy (>0.5-2.0 years), (C) early childhood (>2.0-5.0 years), (D) mid-childhood (>5.0-9.0 years), (E) late childhood (>9.0-14.0 years), (F) adolescence (>14.0-19.0 years) Models are adjusted for sex of child, maternal age at child’s birth, maternal education, maternal height, maternal pre-pregnancy BMI, maternal smoking during pregnancy, parity, maternal ethnic background, gestational diabetes, gestational hypertension, pre-eclampsia. Specific variables were not available in few cohorts (see Table 1), hence not adjusted for.

**S8 Fig.**
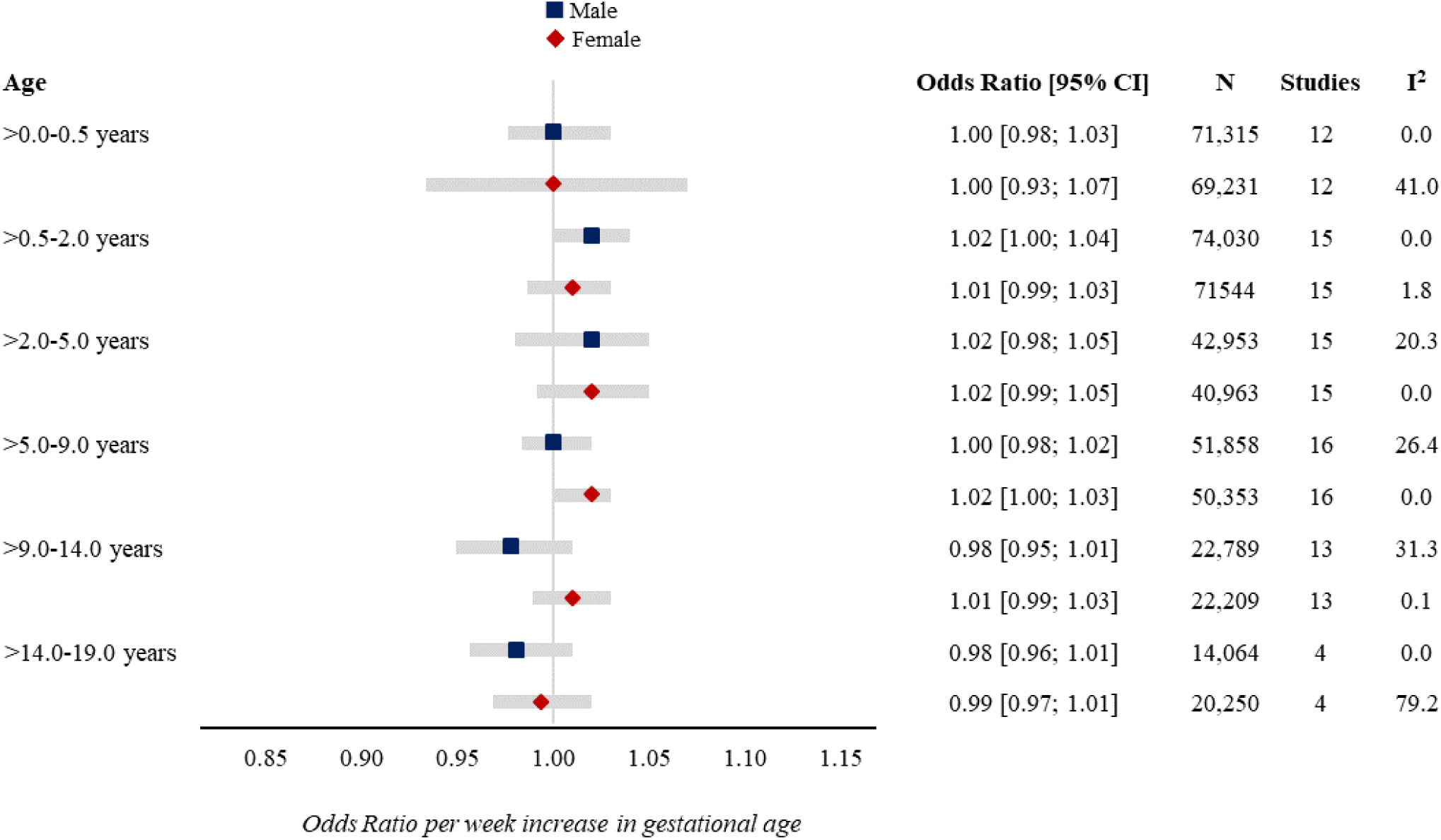
Forest plot of associations between GA (completed weeks) and odds of overweight by sex. Estimates from study-specific logistic regression models were assigned weights under the random-effects model to attain overall estimates. Results reflect odds ratio of overweight (vs. normal weight) per week increase in gestational age at birth in early infancy (>0.0-0.5 years), late infancy (>0.5-2.0 years), early childhood (>2.0-5.0 years), mid-childhood (>5.0-9.0 years), late childhood (>9.0-14.0 years), adolescence (>14.0-19.0 years). Models are adjusted for sex of child, maternal age at child’s birth, maternal education, maternal height, maternal pre-pregnancy BMI, maternal smoking during pregnancy, parity, maternal ethnic background, gestational diabetes, gestational hypertension, pre-eclampsia. Specific variables were not available in few cohorts (see Table 1), hence not adjusted for.

**S9 Fig.**
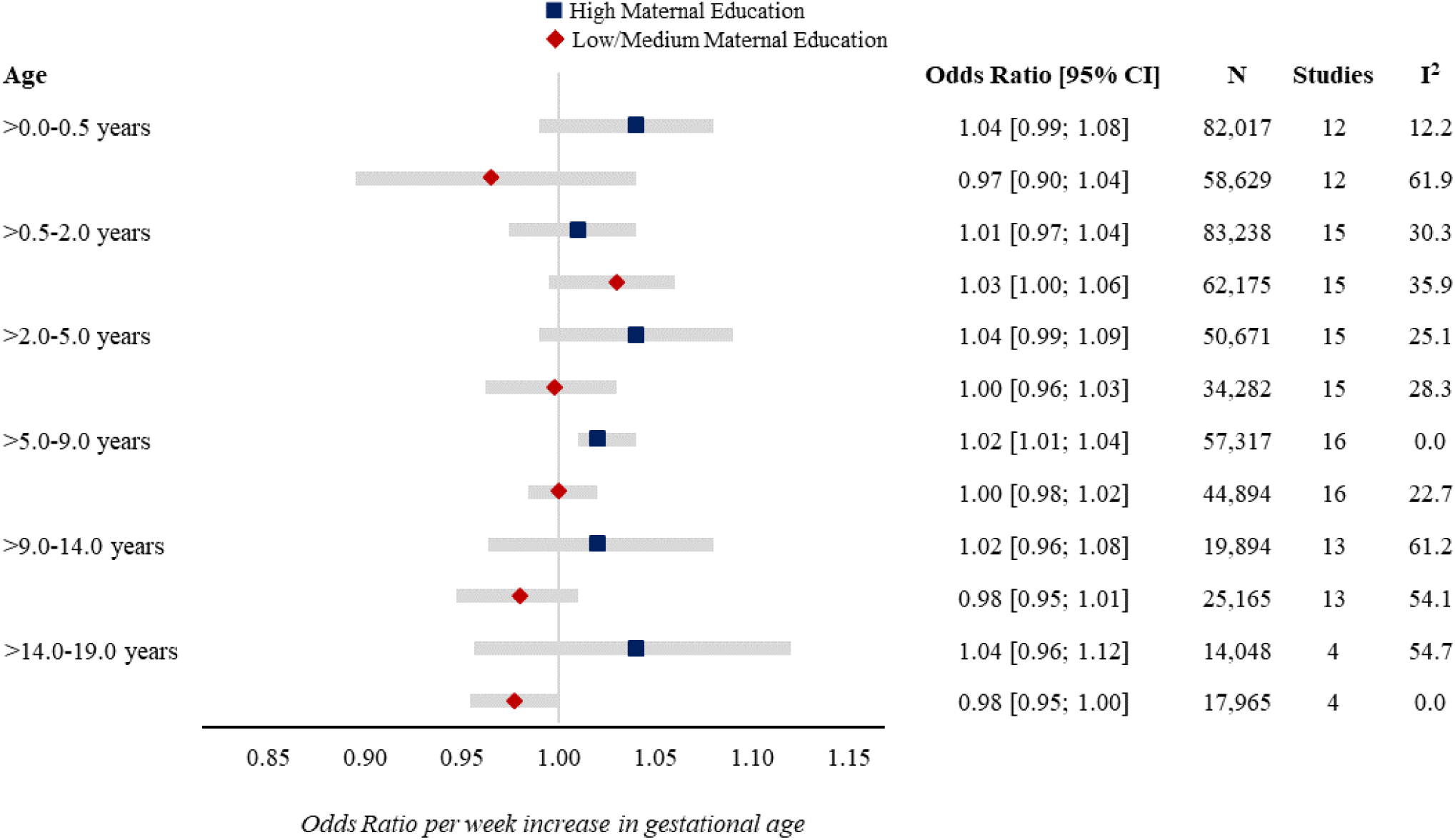
Forest plot of associations between GA (completed weeks) and odds of overweight by maternal education. Estimates from study-specific logistic regression models were assigned weights under the random-effects model to attain overall estimates. Results reflect odds ratio of overweight (vs. normal weight) per week increase in gestational age at birth in early infancy (>0.0-0.5 years), late infancy (>0.5-2.0 years), early childhood (>2.0-5.0 years), mid-childhood (>5.0-9.0 years), late childhood (>9.0-14.0 years), adolescence (>14.0-19.0 years). Models are adjusted for sex of child, maternal age at child’s birth, maternal education, maternal height, maternal pre-pregnancy BMI, maternal smoking during pregnancy, parity, maternal ethnic background, gestational diabetes, gestational hypertension, pre-eclampsia. Specific variables were not available in few cohorts (see Table 1), hence not adjusted for.

**S10 Fig.**
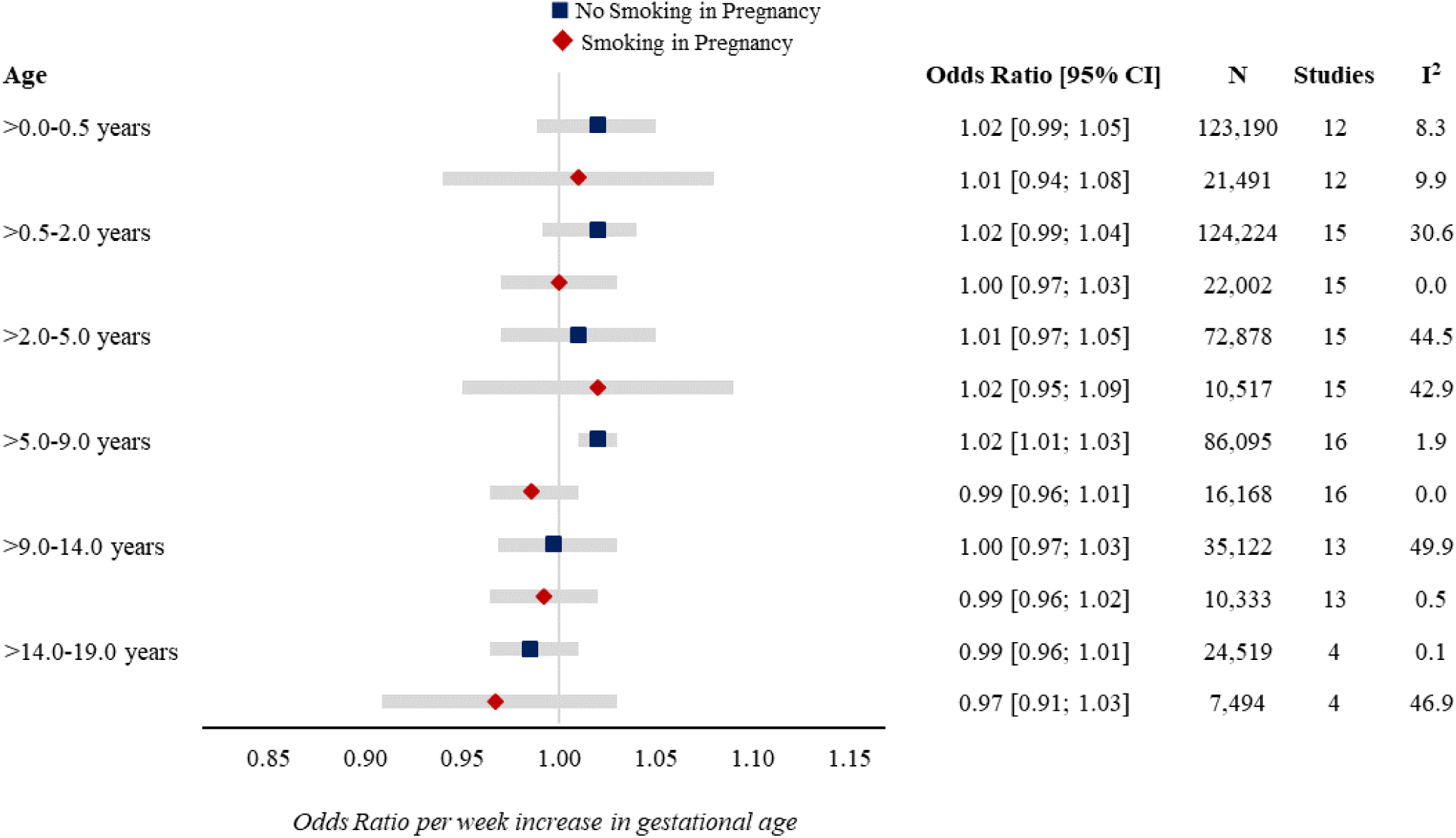
Forest plot of associations between GA (completed weeks) and odds of overweight by maternal smoking in pregnancy. Estimates from study-specific logistic regression models were assigned weights under the random-effects model to attain overall estimates. Results reflect odds ratio of overweight (vs. normal weight) per week increase in gestational age at birth in early infancy (>0.0-0.5 years), late infancy (>0.5-2.0 years), early childhood (>2.0-5.0 years), mid-childhood (>5.0-9.0 years), late childhood (>9.0-14.0 years), adolescence (>14.0-19.0 years). Models are adjusted for sex of child, maternal age at child’s birth, maternal education, maternal height, maternal pre-pregnancy BMI, maternal smoking during pregnancy, parity, maternal ethnic background, gestational diabetes, gestational hypertension, pre-eclampsia. Specific variables were not available in few cohorts (see Table 1), hence not adjusted for.

### S1 Text. Information about variable classification and coding

#### Gestational Age at Birth

Each cohort decided the most accurate available measure of GA, but a priority was given to last menstrual period (LMP), unless it varied from an ultrasound-based (US) estimate by more than 1 week, in which ultrasound was used. If LMP was not available, US was used. If LMP or US was not available, information from a maternal report was used.

#### Maternal Education

Maternal education was classified according to International Classification of Education 97/2011 (ISCED 97/2011) as either *high, medium* or *low.* High includes short cycle tertiary, Bachelor, Masters, Doctoral or equivalent (ISCED-2011: 5-8, ISCED-97: 5-6); Medium includes Upper Secondary, Post-secondary non-tertiary (ISCED-2011: 3-4, ISCED-97: 3-4); Low includes No education, early childhood, pre-primary, primary, lower secondary or second stage of basic education (ISCED-2011: 0-2, ISCED-97: 0-2).

#### Maternal Ethnic Background

Maternal ethnic background was based on (a) *colour of mother* (white/Caucasian, non-white/non-Caucasian) or (b) *country of origin of parents* (western, non-western or mixed), where western countries include European Union, Andorra, Australia, Canada, Iceland, Liechtenstein, Monaco, New Zealand, Norway, San Marino, Switzerland, USA and Vatican City. Non-western countries include all other countries, while mixed refers to one parent from a western country and one parent from a non-western country. The distinction between and definition of western and non-western is chosen in accordance with Statistics Denmark (https://www.dst.dk/Site/Dst/SingleFiles/GetArchiveFile.aspx?fi=91448101625&fo=0&ext=kvaldel, guidelines, which is a continuation of a former UN-definition

### S1 Appendix. Cohort-specific sources of funding/support

#### ALSPAC

Core funding for the Avon Longitudinal Study of Parents and Children (ALSPAC) is provided by the UK Medical Research Council and Wellcome (217065/Z/19/Z) and the University of Bristol. A comprehensive list of grants funding is available on the ALSPAC website (http://www.bristol.ac.uk/alspac/external/documents/grant-acknowledgements.pdf). DAL and AK work in a unit that is supported by the University of Bristol and UK Medical Research Council (MC_UU_00011/6) and DAL holds a European Research Council Advanced Grant (ERC grant agreement no 669545) and is a NIHR Senior Investigator (NF-0616-10102). The funders had no role in the design of the study, the collection, analysis, or interpretation of the data; the writing of the manuscript, or the decision to submit the manuscript for publication. The views expressed in this paper are those of the authors and not necessarily those of any funder.

#### AOF

All Our Families is funded through Alberta Innovates Interdisciplinary Team Grant #200700595, the Alberta Children’s Hospital Foundation, the Canadian Institutes of Health Research, and the Social Sciences and Humanities Research Council. Initial funding to investigate maternal and infant outcomes during the perinatal period was provided by Three Cheers for the Early Years, Alberta Health Services. Expansion of the cohort, to include collection of biological specimens to predict determinants of adverse birth outcomes, was funded by Alberta Innovates Health Solutions [Interdisciplinary Team Grant #200700595] and the Global Alliance to Prevent Prematurity and Stillbirth [GAPPS award #12006]. Subsequent collaborations focusing on the developmental trajectories of AOB children have been supported by numerous grants, including the Social Sciences and Humanities Research Council, Alberta Health Services, the Alberta Children’s Hospital Foundation, the Alberta Centre for Child, Family and Community Research, Upstart the United Way of Calgary, Women and Children’s Health Research Institute, the Canadian Foundation for Fetal Alcohol Research and the Max Bell Foundation.

#### BIB

BiB receives core infrastructure funding from the Wellcome Trust (WT101597MA) and a joint grant from the UK Medical Research Council (MRC) and Economic and Social Science Research Council (ESRC) (MR/N024397/1). This study has received support from the British Heart Foundation (CS/16/4/32482), US National Institutes of Health (R01 DK10324), European Research Council under the European Union’s Seventh Framework Programme (FP7/2007-2013) / ERC grant agreement no 669545, and National Institute for Health Research ARC Yorkshire and Humber (NIHR200166. PMW receives funding from the National Institute for Health Research Applied Research Collaboration for Greater Manchester. The views expressed are those of the author(s), and not necessarily those of the NHS, the NIHR or the Department of Health and Social Care.

#### CHILD

The CHILD Cohort Study was launched in 2008 with funding from AllerGen NCE Inc. and the Canadian Institutes of Health Research (CIHR). Additional funding has been provided for the core study by Genome Canada and local provincial funding and philanthropic support. The funders had no role in the design of the study, the collection, analysis, or interpretation of the data; the writing of the manuscript, or the decision to submit the manuscript for publication. The views expressed in this paper are those of the authors and not necessarily those of any funder

#### DNBC

The Danish National Birth Cohort was established with a significant grant from the Danish National Research Foundation. Additional support was obtained from the Danish Regional Committees, the Pharmacy Foundation, the Egmont Foundation, the March of Dimes Birth Defects Foundation, the Health Foundation and other minor grants. The DNBC Biobank has been supported by the Novo Nordisk Foundation and the Lundbeck Foundation. Follow-up of mothers and children have been supported by the Danish Medical Research Council (SSVF 0646, 271-08-0839/06-066023, O602-01042B, 0602-02738B), the Lundbeck Foundation (195/04, R100-A9193), The Innovation Fund Denmark 0603-00294B (09–067124), the Nordea Foundation (02-2013-2014), Aarhus Ideas (AU R9-A959-13-S804), University of Copenhagen Strategic Grant (IFSV 2012), and the Danish Council for Independent Research (DFF – 4183-00594 and DFF - 4183-00152). AP is funded by a Lundbeck Foundation fellowship (R264-2017-3099)

#### EDEN

The EDEN study was supported by Foundation for medical research (FRM), National Agency for Research (ANR), National Institute for Research in Public health (IRESP: TGIR cohorte santé 2008 program), French Ministry of Health (DGS), French Ministry of Research, INSERM Bone and Joint

Diseases National Research (PRO-A) and Human Nutrition National Research Programs, Paris-Sud University, Nestlé, French National Institute for Population Health Surveillance (InVS), French National Institute for Health Education (INPES), the European Union FP7 programmes (FP7/2007-2013, HELIX, ESCAPE, ENRIECO, Medall projects), Diabetes National Research Program (through a collaboration with the French Association of Diabetic Patients (AFD)), French Agency for Environmental Health Safety (now ANSES), Mutuelle Générale de l’Education Nationale a complementary health insurance (MGEN), French national agency for food security, French speaking association for the study of diabetes and metabolism (ALFEDIAM).

#### ELFE

The Elfe cohort received funding from the National Research Agency Investment for the Future program [ANR-11-EQPX-0038]; French National Institute for Research in Public Health (IRESP TGIR 2009-2001 program); Ministry of Higher Education and Research; Ministry of Environment; Ministry of Health; French Agency for Public Health; Ministry of Culture; and National Family Allowance Fund.

#### G21

Generation XXI was supported by the European Regional Development Fund (ERDF) through the Operational Programme Competitiveness and Internationalization and national funding from the Foundation for Science and Technology (FCT), Portuguese Ministry of Science, Technology and Higher Education under the project “HIneC: When do health inequalities start? Understanding the impact of childhood social adversity on health trajectories from birth to early adolescence” (POCI-01-0145-FEDER-029567; Reference PTDC/SAU-PUB/29567/2017). It is also supported by the Unidade de Investigação em Epidemiologia - Instituto de Saúde Pública da Universidade do Porto (EPIUnit) (UIDB/04750/2020), Administração Regional de Saúde Norte (Regional Department of Ministry of Health) and Fundação Calouste Gulbenkian; PhD Grant SFRH/BD/108742/2015 (to SS) co-funded by FCT and the Human Capital Operational Programme (POCH/FSE Program). This project received funding from the European Union’s Horizon 2020 research and innovation programme under the project ; EUCAN-Connect grant agreement No 824989. ACS is founded by a FCT Investigator contracts IF/01060/2015.

#### GECKO

The GECKO Drenthe birth cohort was funded by an unrestricted grant of Hutchison Whampoa Ltd, Hong Kong and supported by the University of Groningen, Well Baby Clinic Foundation Icare, Noordlease, Paediatric Association Of The Netherlands, Youth Preventive Health Care Drenthe and European Union’s Horizon 2020 research and innovation programme (LifeCycle, Grant Agreement No. 733206 LifeCycle).

#### Generation R

The general design of the Generation R Study is made possible by financial support from the Erasmus MC, University Medical Center, Rotterdam, Erasmus University Rotterdam, Netherlands Organization for Health Research and Development (ZonMw), Netherlands Organisation for Scientific Research (NWO), Ministry of Health, Welfare and Sport and Ministry of Youth and Families. This project received funding from the European Union’s Horizon 2020 research and innovation programme (LIFECYCLE, grant agreement No 733206, 2016; EUCAN-Connect grant agreement No 824989; ATHLETE, grant agreement No 874583). VJ received funding from a Consolidator Grant from the European Research Council (ERC-2014-CoG-648916). The study sponsors had no role in the study design, data analysis, interpretation of data, or writing of this report.

#### INMA

INMA-Valencia was funded by Grants from UE (FP7-ENV-2011 cod 282957 and HEALTH.2010.2.4.5-1), Spain: ISCIII (G03/176; FIS-FEDER: PI09/02647, PI11/01007, PI11/02591, PI11/02038, PI13/1944, PI13/2032, PI14/00891, PI14/01687, and PI16/1288; Miguel Servet-FEDER CP11/00178, CP15/00025, and CPII16/00051), and Generalitat Valenciana: FISABIO (UGP 15-230, UGP-15-244,and UGP-15-249). INMA-Gipuzkoa was funded by grants from the Instituto de Salud Carlos III (FISFIS PI06/0867, FISPS09/0009) 0867,Red INMA G03/176) and the Departamento de Salud del Gobierno Vasco (2005111093 and 2009111069) and the Provincial Government of Guipúzcoa (DFG06/004 and FG08/001). INMA-Sabadell was funded by grants from ISCIII (Red INMA G03/176; CB06/02/0041; FIS-FEDER: PI041436; PI081151; PI12/01890; CP13/00054; PI15/00118; CP16/00128; PI16/00118; PI16/00261; PI18/00547), CIBERESP, Generalitat de Catalunya-CIRIT (1999SGR 00241), Generalitat de Catalunya-AGAUR (2009 SGR 501, 2014 SGR 822), Fundació La Marató de TV3 (090430), Spanish Ministry of Economy and Competitiveness (SAF2012-32991 incl. FEDER funds), Agence Nationale de Securite Sanitaire de l’Alimentation de l’Environnement et du Travail (1262C0010; EST-2016 RF-21), and the European Commission (261357, 308333, 603794 and 634453). ISGlobal acknowledges support from the Spanih Ministry of Science, Innovation and Universities through the “Centro de Excelencia Severo Ochoa 2019-2023” Program (CEX2018-000806-S), and support from the Generalitat de Catalunya through the CERCA Program.

#### MoBa

The Norwegian Mother, Father and Child Cohort Study is supported by the Norwegian Ministry of Health and Care Services and the Ministry of Education and Research. JN received funding, in part, from the European Union’s Horizon 2020 research and innovation programme (LIFECYCLE, grant agreement No 733206), The work of JRH was partly supported by the Research Council of Norway through its Centres of Excellence funding scheme, project number 262700.

#### NINFEA

The NINFEA cohort was initially funded by the Compagnia SanPaolo Foundation and the Piedmont Region. It received funding from European projects: CHICOS (FP7 grant number HEALTH-FP7-2009-241604, LifeCycle (H2020 grant number 733206), ATHLETE (H2020 grant number 874583).

#### NFBC1986

NFBC1986 received financial support from EU QLG1-CT-2000-01643 (EUROBLCS, grant number E51560), NorFA (grant numbers 731, 20056 and 30167) and USA / NIH 2000 G DF682 (grant number 50945). Financial support for data generation, research and supporting staff was received from the Academy of Finland (grant numbers: 104781, 120315, 129269, 1114194, 24300796, 285547 (EGEA)); University Hospital Oulu, Biocenter, University of Oulu, Finland (grant number: 75617); NIHM (grant number: MH063706, Smalley and Jarvelin for NFBC1986 data collection), Juselius Foundation; NIH/NIMH (grant number: 5R01MH63706:02); the European Commission: EURO-BLCS, Framework 5 award QLG1-CT-2000-01643 (for NFBC1986 data collection), ENGAGE project and grant agreement HEALTH-F4-2007 (grant number: 201413); EU H2020-HCO-2004 iHEALTH Action (grant number: 643774), EU H2020-PHC-2014 DynaHealth Action (grant number: 633595); ALEC Action (grant number: 633212); ERDF European Regional Development Fund (grant number: 539/2010 A31592); the Medical Research Council (MRC), UK (grant numbers: G0500539, G0600705, G1002319, MR/M013138/1), EU H2020-SC1-2016-2017 LifeCycle Action (grant number: 733206). The programme is currently funded by EU H2020-SC1-2016-2017 LifeCycle Action (grant number: 733206), EU-H2020 EUCAN Connect (grant number: 824989) and H2020-SC1-2019 LongI-Tools (grant number 874739).

#### The Raine study

The Raine study has been funded by program and project grants from the Australian National Health and Medical Research Council (NHMRC), the Commonwealth Scientific and Industrial Research Organisation, Healthway and the Lions Eye Institute in Western Australia. The Raine study Gen2-17 year follow-up was funded by the NHMRC Program Grant (Stanley et al, ID 353514). The Raine study participation in LIFECYCLE was funded by a grant from the National Health and Medical Research Council, Australia (GNT114285). The University of Western Australia (UWA), Curtin University, the Raine Medical Research Foundation, the Telethon Kids Institute, the Women’s and Infant’s Research Foundation (KEMH), Murdoch University, The University of Notre Dame Australia and Edith Cowan University provide funding for the Core Management of the Raine Study. RCH was supported by NHMRC fellowship (Grant Number 1053384). JC is supported by NHMRC EU (Grant Number 114285).

#### SWS

The SWS is supported by grants from the Medical Research Council, National Institute for Health Research Southampton Biomedical Research Centre, British Heart Foundation, University of Southampton and University Hospital Southampton National Health Service Foundation Trust, and the European Union’s Seventh Framework Programme (FP7/2007-2013), project Early Nutrition (grant 289346) and from the European Union’s Horizon 2020 research and innovation programme (LIFECYCLE, grant agreement No 733206). Study participants were drawn from a cohort study funded by the Medical Research Council and the Dunhill Medical Trust.

### S2 Appendix. Cohort-specific acknowledgments

#### ALSPAC

We are extremely grateful to all of the families who took part in ALSPAC, the midwives for their help in recruiting them, and the whole ALSPAC team, which includes interviewers, computer and laboratory technicians, clerical workers, research scientists, volunteers, managers, receptionists and nurses.

#### AOF

The authors acknowledge the tremendous contribution and support of AOF participants and AOF team members. We are extremely grateful to the investigators, coordinators, research assistants, graduate and undergraduate students, volunteers, clerical staff and managers.

#### BIB

The authors acknowledge that Born in Bradford is only possible because of the enthusiasm and commitment of the children and parents in Born in Bradford. We are grateful to all participants, health professionals and researchers who have made Born in Bradford happen.

#### CHILD

We thank the CHILD Cohort Study (CHILD) participant families for their dedication and commitment to advancing health research. Visit CHILD at childcohort.ca.

#### DNBC

The authors would like to thank the participants, the first Principal Investigator of DNBC Prof. Jørn Olsen, the scientific managerial team, and DNBC secretariat for being, establishing, developing and consolidating the Danish National Birth Cohort.

#### EDEN

The authors thank the cohort participants and the EDEN mother-child study group, whose members are: I. Annesi-Maesano, J.Y. Bernard, J. Botton, M.A. Charles, P. Dargent-Molina, B. de Lauzon-Guillain, P. Ducimetière, M. de Agostini, B. Foliguet, A. Forhan, X. Fritel, A. Germa, V. Goua, R. Hankard, B. Heude, M. Kaminski, B. Larroque†, N. Lelong, J. Lepeule, G. Magnin, L. Marchand, C. Nabet, F Pierre, R. Slama, M.J. Saurel-Cubizolles, M. Schweitzer, O. Thiebaugeorges.

#### ELFE

The authors are grateful to 1) the former members of the Elfe unit without whom the project would never have started: Henri Léridon, initiator and former Principal Investigator of the project, Stéphanie Vandentorren, Claudine Pirus, and Ando Rakotonirina; 2) the expertise and assistance of members of the unit for support functions, 3) all the researchers who contribute to the projects as members of the Elfe thematic groups and especially their coordinators; 4) all the field research assistants and interviewers; 5) and above all, all the Elfe families who have placed their confidence in us and given up their time to the study

#### G21

The authors gratefully acknowledge the families enrolled in Generation XXI for their kindness, all members of the research team for their enthusiasm and perseverance, and the participating hospitals and their staff for their help and support.

#### GECKO

The authors are grateful to the families who took part in the GECKO Drenthe study, the midwives, gyneacologists, nurses, and the general practitioners and all health professionals at the Preventive Child Healthcare Drenthe for their help in the recruitment and the measurements, and the GECKO Drenthe study team.

#### Generation R

The authors gratefully acknowledge the contribution of participants, research collaborators, general practitioners, hospitals, midwives, and pharmacies in Rotterdam.

#### INMA

The authors would particularly like to thank all the participants for their generous collaboration. The authors are grateful to Silvia Fochs, Nuria Pey, Mireia Garcia, Maria Victoria Estraña, Maria Victoria Iturriaga, Cristina Capo and Josep LLuch for their assistance in contacting the families and administering the questionnaires.

#### MoBa

The authors are grateful to all the participating families in Norway who take part in this on-going cohort study.

#### NFBC1986

The authors thank all cohort members and researchers who have participated in the NFBC studies. We also wish acknowledge the work of the NFBC project center.

#### NINFEA

The authors thank all families participating in the NINFEA cohort.

#### The Raine study

The authors would like to acknowledge the Raine study participants and their families for their ongoing participation in the study and the Raine study team for study co-ordination and data collection. We also thank the NHMRC for their long term contribution to funding the study over the last 30 years. The core management of the Raine study is funded by The University of Western Australia, Curtin University, Telethon Kids Institute, Women and Infants Research Foundation, Edith Cowan University, Murdoch University, The University of Notre Dame Australia and the Raine Medical Research Foundation.

#### SWS

The authors are grateful to the women and their children in Southampton who gave their time to take part in the Southampton Women’s Survey and to the research nurses and other staff who collected and processed the data.

### S3 Appendix. Cohort-specific ethics approval

#### ALSPAC

Ethical approval for the study was obtained from the ALSPAC Ethics and Law Committee and the Local Research Ethics Committees

#### AOF

The All Our Families study was approved by the Child Health Research Office and the Conjoint Health Research Ethics Board of the Faculties of Medicine, Nursing, and Kinesiology, University of Calgary, and the Affiliated Teaching Institutions (Ethics ID 20821 and 22821).

#### BiB

Ethics approval has been obtained for the main platform study and all of the individual substudies from the Bradford Research Ethics Committee

#### CHILD

Ethics approvals for the study were obtained at recruitment and at each data collection phase from all four Canadian sites.

#### DNBC

The DNBC complies with the Declaration of Helsinki and was approved by the Danish National Committee on Biomedical Research Ethics

#### EDEN

The study received approval from the ethics committee (CCPPRB) of Kremlin Bicêtre on 12 December 2002 and from CNIL (Commission Nationale Informatique et Liberté), the French data privacy institution

#### ELFE

Ethical approvals for data collection in maternity units and for each data collection wave during follow-up were obtained from the national advisory committee on information processing in health research (CCTIRS: Comité Consultatif sur le Traitement de l’Information en matière de Recherche dans le domaine de la Santé), the national data protection authority (CNIL: Comission Nationale Informatique et Liberté) and, in case of invasive data collection such as biological sampling, the committee for protection of persons engaged in research (CPP: Comité de Protection des Personnes). The ELFE study was also approved by the national committee for statistical information (CNIS: Conseil National de l’Information Statistique).

#### G21

Generation XXI was approved by the Portuguese Data Protection Authority and by the Ethics Committee of Hospital São João, and data confidentiality and protection were guaranteed in all procedures according to the Declaration of Helsinki. Signed informed consent was obtained for all adults and children participants had it signed by their legal guardian at every study waves.

#### GECKO

The GECKO Drenthe study complies with the Declaration of Helsinki and was approved by the Medical Ethics Committee of the University Medical Center Groningen (UMCG).

#### Generation R

The general design, all research aims and the specific measurements in the Generation R Study have been approved by the Medical Ethical Committee of the Erasmus Medical Center, Rotterdam. New measurements will only be embedded in the study after approval of the Medical Ethical Committee.

#### INMA

The INMA project was approved by the ethics committee in each area.

#### MoBa

The establishment and data collection in MoBa was previously based on a license from the Norwegian Data protection agency and approval from The Regional Committee for Medical Research Ethics, and it is now based on regulations related to the Norwegian Health Registry Act.

#### NINFEA

The Ethical Committee of the San Giovanni Battista Hospital and CTO/CRF/Maria Adelaide Hospital of Turin approved the NINFEA study (approval N. 0048362, and subsequent amendments)

#### NFBC1986

The study was performed according to the Declaration of Helsinki, and approved by the Ethics committee of the Northern Ostrobothnia Hospital District in Finland

#### The Raine study

The original cohort study received approval from the King Edward Memorial Hospital for women ethics committee in 1989 (DD/JS/459), and all subsequent follow-ups also received institutional human research ethics committee (HREC) approval prior to commencing. All participants were provided with participant information sheets and parents (Gen1) provided informed consent, and the child (Gen2) provided assent. When the Raine study Gen2 participants turned 18 years of age, ethics approval was further received from the University of Western Australia HREC (RA/4/1/2100) to contact and obtain consent from Raine study participants for any data that was collected before they were 18 to be used for future research. A further UWA HREC was provided to all a single ‘overarching’ approval code that recognises all previous approvals under which previous data and/or bio-samples were collected. This approval was received on 29 April 2020 and provides a single consolidated approval (RA/4/20/5722) for use of research data and/or bio-samples held in the Raine study data collection.

#### SWS

The study had full approval at each wave from the Southampton and Southwest Hampshire Local Research Ethics Committee.

### S4 Appendix. Cohort-specific consent to participate

#### ALSPAC

Informed consent for the use of data collected via questionnaires and clinics was obtained from participants following the recommendations of the ALSPAC Ethics and Law Committee at the time.

#### AOF

Participants provided informed consent at the time of recruitment and were provided copies of the consent form for their records.

#### BiB

All participants gave written informed consent.

#### CHILD

Informed consent of CHILD Cohort Study participants was obtained upon enrolment and at each subsequent data collection phase.

#### DNBC

Informed consent was obtained from participants upon enrolment

#### EDEN

Women gave written informed consent for themselves and their child. Fathers gave written informed consent for themselves.

#### ELFE

Informed consent was signed by the parents or the mother alone, with the father being informed of his right to deny consent for participation

#### G21

Written informed consent was signed by any of the parents or legal representative of each participant at the baseline and in all subsequent follow up evaluations. Even with consent of the parents, when the child is not willing to participate, no measurements were performed. From the age of 13 years, children were also asked for their informed consent.

#### GECKO

Parents of all participants in the study gave written informed consent.

#### Gen R

Participants are asked for their written informed consent for the four consecutive phases of the study (prenatally, birth to 4 years, 4–12 years, and from 12 years onwards). At the start of each phase, mothers and their partners receive written and oral information about the study. Even with consent of the parents, when the child is not willing to participate actively, no measurements are performed. From the age of 12 years, children are asked for written informed consent

#### INMA

All participants provided written informed consent before enrolment to the study

#### MoBa

MoBa is conducted according to the guidelines laid down in the declaration of Helsinki, and written informed consent was obtained from all participants. A detailed protocol of the study including the consent can be found elsewhere (http://www.fhi.no/morogbarn).

#### NINFEA

Informed consent was obtained from all the participants at enrolment and at each follow-up.

#### NFBC1986

Each participant and a guardian gave signed informed consent

#### The Raine study

All participants were provided with participant information sheets and parents (Gen1) provided informed consent, and the child (Gen2) provided assent. When the Raine study Gen2 participants turned 18 years of age, ethics approval was further received from the University of Western Australia HREC (RA/4/1/2100) to contact and obtain consent from Raine study participants for any data that was collected before they were 18 to be used for future research.

#### SWS

All participants gave written informed consent for each data collection wave of the study.

## References

1. Goldenberg RL, Culhane JF, Iams JD, Romero R. Epidemiology and causes of preterm birth. Lancet. 2008;371(9606):75–84.

2. Saigal S, Doyle LW. An overview of mortality and sequelae of preterm birth from infancy to adulthood. Lancet. 2008;371(9608):261–9.

3. Bick D. Born too soon: the global issue of preterm birth. Midwifery. 2012;28(4):341–2.

4. Chawanpaiboon S, Vogel JP, Moller AB, Lumbiganon P, Petzold M, Hogan D, et al. Global, regional, and national estimates of levels of preterm birth in 2014: a systematic review and modelling analysis. Lancet Glob Health. 2019;7(1):e37–e46.

5. Markopoulou P, Papanikolaou E, Analytis A, Zoumakis E, Siahanidou T. Preterm Birth as a Risk Factor for Metabolic Syndrome and Cardiovascular Disease in Adult Life: A Systematic Review and Meta-Analysis. J Pediatr. 2019;210:69–80 e5.

6. Parkinson JR, Hyde MJ, Gale C, Santhakumaran S, Modi N. Preterm birth and the metabolic syndrome in adult life: a systematic review and meta-analysis. Pediatrics. 2013;131(4):e1240–63.

7. Kajantie E, Strang-Karlsson S, Evensen KAI, Haaramo P. Adult outcomes of being born late preterm or early term - What do we know? Semin Fetal Neonatal Med. 2019;24(1):66–83.

8. Johansson S, Iliadou A, Bergvall N, Tuvemo T, Norman M, Cnattingius S. Risk of high blood pressure among young men increases with the degree of immaturity at birth. Circulation. 2005;112(22):3430–6.

9. Lawlor DA, Hubinette A, Tynelius P, Leon DA, Smith GD, Rasmussen F. Associations of gestational age and intrauterine growth with systolic blood pressure in a family-based study of 386,485 men in 331,089 families. Circulation. 2007;115(5):562–8.

10. Sipola-Leppanen M, Vaarasmaki M, Tikanmaki M, Matinolli HM, Miettola S, Hovi P, et al. Cardiometabolic risk factors in young adults who were born preterm. Am J Epidemiol. 2015;181(11):861–73.

11. Sipola-Leppanen M, Vaarasmaki M, Tikanmaki M, Hovi P, Miettola S, Ruokonen A, et al. Cardiovascular risk factors in adolescents born preterm. Pediatrics. 2014;134(4):e1072–81.

12. Baird J, Jacob C, Barker M, Fall CH, Hanson M, Harvey NC, et al. Developmental Origins of Health and Disease: A Lifecourse Approach to the Prevention of Non-Communicable Diseases. Healthcare (Basel). 2017;5(1).

13. Gluckman PD, Hanson MA, Cooper C, Thornburg KL. Effect of in utero and early-life conditions on adult health and disease. N Engl J Med. 2008;359(1):61–73.

14. Hanson MA, Gluckman PD. Early developmental conditioning of later health and disease: physiology or pathophysiology? Physiol Rev. 2014;94(4):1027–76.

15. Kopec G, Shekhawat PS, Mhanna MJ. Prevalence of diabetes and obesity in association with prematurity and growth restriction. Diabetes Metab Syndr Obes. 2017;10:285–95.

16. Remacle C, Bieswal F, Reusens B. Programming of obesity and cardiovascular disease. Int J Obes Relat Metab Disord. 2004;28 Suppl 3:S46–53.

17. Rowe DL, Derraik JG, Robinson E, Cutfield WS, Hofman PL. Preterm birth and the endocrine regulation of growth in childhood and adolescence. Clin Endocrinol (Oxf). 2011;75(5):661–5.

18. Barker DJ. Obesity and early life. Obes Rev. 2007;8 Suppl 1:45–9.

19. Barker DJ. Adult consequences of fetal growth restriction. Clin Obstet Gynecol. 2006;49(2):270–83.

20. Euser AM, de Wit CC, Finken MJ, Rijken M, Wit JM. Growth of preterm born children. Horm Res. 2008;70(6):319–28.

21. Griffin IJ, Cooke RJ. Development of whole body adiposity in preterm infants. Early Hum Dev. 2012;88 Suppl 1:S19–24.

22. WHO European Regional Obesity Report 2022. Copenhagen; 2022 2022.

23. Hollanders JJ, van der Pal SM, van Dommelen P, Rotteveel J, Finken MJJ. Growth pattern and final height of very preterm vs. very low birth weight infants. Pediatr Res. 2017;82(2):317–23.

24. Adams M, Andersen AM, Andersen PK, Haig D, Henriksen TB, Hertz-Picciotto I, et al. Sostrup statement on low birthweight. Int J Epidemiol. 2003;32(5):884–5.

25. Ananth CV, Schisterman EF. Confounding, causality, and confusion: the role of intermediate variables in interpreting observational studies in obstetrics. Am J Obstet Gynecol. 2017;217(2):167–75.

26. Roberts G, Cheong J, Opie G, Carse E, Davis N, Duff J, et al. Growth of extremely preterm survivors from birth to 18 years of age compared with term controls. Pediatrics. 2013;131(2):e439–45.

27. Roswall J, Karlsson AK, Allvin K, Tangen GA, Bergman S, Niklasson A, et al. Preschool children born moderately preterm have increased waist circumference at two years of age despite low body mass index. Acta Paediatr. 2012;101(11):1175–81.

28. Bortolotto CC, Santos IS, Dos Santos Vaz J, Matijasevich A, Barros AJD, Barros FC, et al. Prematurity and body composition at 6, 18, and 30 years of age: Pelotas (Brazil) 2004, 1993, and 1982 birth cohorts. BMC Public Health. 2021;21(1):321.

29. Mardones F, Villarroel L, Karzulovic L, Barja S, Arnaiz P, Taibo M, et al. Association of perinatal factors and obesity in 6- to 8-year-old Chilean children. Int J Epidemiol. 2008;37(4):902–10.

30. Massion S, Wickham S, Pearce A, Barr B, Law C, Taylor-Robinson D. Exploring the impact of early life factors on inequalities in risk of overweight in UK children: findings from the UK Millennium Cohort Study. Arch Dis Child. 2016;101(8):724–30.

31. Ni Y, Beckmann J, Gandhi R, Hurst JR, Morris JK, Marlow N. Growth to early adulthood following extremely preterm birth: the EPICure study. Arch Dis Child Fetal Neonatal Ed. 2020;105(5):496–503.

32. Bocca-Tjeertes IF, Kerstjens JM, Reijneveld SA, de Winter AF, Bos AF. Growth and predictors of growth restraint in moderately preterm children aged 0 to 4 years. Pediatrics. 2011;128(5):e1187–94.

33. Dietz WH. Critical periods in childhood for the development of obesity. Am J Clin Nutr. 1994;59(5):955–9.

34. Cooper R, Atherton K, Power C. Gestational age and risk factors for cardiovascular disease: evidence from the 1958 British birth cohort followed to mid-life. Int J Epidemiol. 2009;38(1):235–44.

35. Andraweera PH, Condon B, Collett G, Gentilcore S, Lassi ZS. Cardiovascular risk factors in those born preterm - systematic review and meta-analysis. J Dev Orig Health Dis. 2021;12(4):539–54.

36. Yoshida-Montezuma Y, Stone E, Iftikhar S, De Rubeis V, Andreacchi AT, Keown-Stoneman C, et al. The association between late preterm birth and cardiometabolic conditions across the life course: A systematic review and meta-analysis. Paediatr Perinat Epidemiol. 2021.

37. Lurbe E, Ingelfinger J. Developmental and Early Life Origins of Cardiometabolic Risk Factors: Novel Findings and Implications. Hypertension. 2021;77(2):308–18.

38. Jaddoe VWV, Felix JF, Andersen AN, Charles MA, Chatzi L, Corpeleijn E, et al. The LifeCycle Project-EU Child Cohort Network: a federated analysis infrastructure and harmonized data of more than 250,000 children and parents. Eur J Epidemiol. 2020;35(7):709–24.

39. Pinot de Moira A, Haakma S, Strandberg-Larsen K, van Enckevort E, Kooijman M, Cadman T, et al. The EU Child Cohort Network’s core data: establishing a set of findable, accessible, interoperable and re-usable (FAIR) variables. Eur J Epidemiol. 2021;36(5):565–80.

40. EUCAN-Connect. A federated FAIR platform enabling large-scale analysis of high-value cohort data connecting Europe and Canada in personalized health [Available from: https://eucanconnect.com/.

41. Fraser A, Macdonald-Wallis C, Tilling K, Boyd A, Golding J, Davey Smith G, et al. Cohort Profile: the Avon Longitudinal Study of Parents and Children: ALSPAC mothers cohort. Int J Epidemiol. 2013;42(1):97–110.

42. Tough SC, McDonald SW, Collisson BA, Graham SA, Kehler H, Kingston D, et al. Cohort Profile: The All Our Babies pregnancy cohort (AOB). Int J Epidemiol. 2017;46(5):1389–90k.

43. Wright J, Small N, Raynor P, Tuffnell D, Bhopal R, Cameron N, et al. Cohort Profile: the Born in Bradford multi-ethnic family cohort study. Int J Epidemiol. 2013;42(4):978–91.

44. Subbarao P, Anand SS, Becker AB, Befus AD, Brauer M, Brook JR, et al. The Canadian Healthy Infant Longitudinal Development (CHILD) Study: examining developmental origins of allergy and asthma. Thorax. 2015;70(10):998–1000.

45. Olsen J, Melbye M, Olsen SF, Sorensen TI, Aaby P, Andersen AM, et al. The Danish National Birth Cohort--its background, structure and aim. Scand J Public Health. 2001;29(4):300–7.

46. Heude B, Forhan A, Slama R, Douhaud L, Bedel S, Saurel-Cubizolles MJ, et al. Cohort Profile: The EDEN mother-child cohort on the prenatal and early postnatal determinants of child health and development. Int J Epidemiol. 2016;45(2):353–63.

47. Charles MA, Thierry X, Lanoe JL, Bois C, Dufourg MN, Popa R, et al. Cohort Profile: The French national cohort of children (ELFE): birth to 5 years. Int J Epidemiol. 2020;49(2):368–9j.

48. Correia S, Rodrigues T, Barros H. Socioeconomic variations in female fertility impairment: a study in a cohort of Portuguese mothers. BMJ Open. 2014;4(1):e003985.

49. L’Abee C, Sauer PJ, Damen M, Rake JP, Cats H, Stolk RP. Cohort Profile: the GECKO Drenthe study, overweight programming during early childhood. Int J Epidemiol. 2008;37(3):486–9.

50. Jaddoe VW, Mackenbach JP, Moll HA, Steegers EA, Tiemeier H, Verhulst FC, et al. The Generation R Study: Design and cohort profile. Eur J Epidemiol. 2006;21(6):475–84.

51. Guxens M, Ballester F, Espada M, Fernandez MF, Grimalt JO, Ibarluzea J, et al. Cohort Profile: the INMA--INfancia y Medio Ambiente--(Environment and Childhood) Project. Int J Epidemiol. 2012;41(4):930–40.

52. Magnus P, Birke C, Vejrup K, Haugan A, Alsaker E, Daltveit AK, et al. Cohort Profile Update: The Norwegian Mother and Child Cohort Study (MoBa). Int J Epidemiol. 2016;45(2):382–8.

53. Jarvelin MR, Hartikainen-Sorri AL, Rantakallio P. Labour induction policy in hospitals of different levels of specialisation. Br J Obstet Gynaecol. 1993;100(4):310–5.

54. Jarvelin MR, Elliott P, Kleinschmidt I, Martuzzi M, Grundy C, Hartikainen AL, et al. Ecological and individual predictors of birthweight in a northern Finland birth cohort 1986. Paediatr Perinat Epidemiol. 1997;11(3):298–312.

55. University of Oulu: Northern Finland Birth Cohort 1986. University of Oulu.

56. Richiardi L, Baussano I, Vizzini L, Douwes J, Pearce N, Merletti F, et al. Feasibility of recruiting a birth cohort through the Internet: the experience of the NINFEA cohort. Eur J Epidemiol. 2007;22(12):831–7.

57. McKnight CM, Newnham JP, Stanley FJ, Mountain JA, Landau LI, Beilin LJ, et al. Birth of a cohort--the first 20 years of the Raine study. Med J Aust. 2012;197(11):608–10.

58. Inskip HM, Godfrey KM, Robinson SM, Law CM, Barker DJ, Cooper C, et al. Cohort profile: The Southampton Women’s Survey. Int J Epidemiol. 2006;35(1):42–8.

59. Doiron D, Marcon Y, Fortier I, Burton P, Ferretti V. Software Application Profile: Opal and Mica: open-source software solutions for epidemiological data management, harmonization and dissemination. Int J Epidemiol. 2017;46(5):1372–8.

60. Swertz MA, Dijkstra M, Adamusiak T, van der Velde JK, Kanterakis A, Roos ET, et al. The MOLGENIS toolkit: rapid prototyping of biosoftware at the push of a button. BMC Bioinformatics. 2010;11 Suppl 12:S12.

61. Swertz MA, Jansen RC. Beyond standardization: dynamic software infrastructures for systems biology. Nat Rev Genet. 2007;8(3):235–43.

62. van der Velde KJ, Imhann F, Charbon B, Pang C, van Enckevort D, Slofstra M, et al. MOLGENIS research: advanced bioinformatics data software for non-bioinformaticians. Bioinformatics. 2019;35(6):1076–8.

63. Budin-Ljosne I, Burton P, Isaeva J, Gaye A, Turner A, Murtagh MJ, et al. DataSHIELD: an ethically robust solution to multiple-site individual-level data analysis. Public Health Genomics. 2015;18(2):87–96.

64. Gaye A, Marcon Y, Isaeva J, LaFlamme P, Turner A, Jones EM, et al. DataSHIELD: taking the analysis to the data, not the data to the analysis. Int J Epidemiol. 2014;43(6):1929–44.

65. Wilson R.C. BOW, Avraam D., Baker J., Tedds J., Turner A., Murtagh M. and Burton P. . DataSHIELD – moving in new directions and dimensions. Data Science Journal. 2017;16.

66. Gill JV, Boyle EM. Outcomes of infants born near term. Arch Dis Child. 2017;102(2):194–8.

67. Woo JG, Daniels SR. Assessment of Body Mass Index in Infancy: It Is Time to Revise Our Guidelines. J Pediatr. 2019;204:10–1.

68. de Onis M, Lobstein T. Defining obesity risk status in the general childhood population: which cut-offs should we use? Int J Pediatr Obes. 2010;5(6):458–60.

69. de Onis M, Onyango AW, Borghi E, Siyam A, Nishida C, Siekmann J. Development of a WHO growth reference for school-aged children and adolescents. Bull World Health Organ. 2007;85(9):660–7.

70. Statistics: UIf. International Standard Classification of Education ISCED 2011. Montréal; 2012.

71. Burke DL, Ensor J, Riley RD. Meta-analysis using individual participant data: one-stage and two-stage approaches, and why they may differ. Stat Med. 2017;36(5):855–75.

72. Borenstein M, Hedges LV, Higgins JP, Rothstein HR. A basic introduction to fixed-effect and random-effects models for meta-analysis. Res Synth Methods. 2010;1(2):97–111.

73. Higgins JP, Thompson SG, Deeks JJ, Altman DG. Measuring inconsistency in meta-analyses. BMJ. 2003;327(7414):557–60.

74. Hastie T, Tibshirani R, Friedman, J. Model Assessment and Selection. The elements of statistical learning: Data mining, inference, and prediction. 6 ed: New York Springer; 2001. p. 219–60.

75. RC T. R: A language and environment for statistical computing. 2014.

76. Viechtbauer W. Conducting Meta-Analyses in R with the metafor Package. Journal of Statistical Software. 2010;36(3).

77. Fewtrell MS, Lucas A, Cole TJ, Wells JC. Prematurity and reduced body fatness at 8-12 y of age. Am J Clin Nutr. 2004;80(2):436–40.

78. Pico C, Palou A. Perinatal programming of obesity: an introduction to the topic. Front Physiol. 2013;4:255.

79. Ong KK, Dunger DB. Perinatal growth failure: the road to obesity, insulin resistance and cardiovascular disease in adults. Best Pract Res Clin Endocrinol Metab. 2002;16(2):191–207.

80. Ong KK, Loos RJ. Rapid infancy weight gain and subsequent obesity: systematic reviews and hopeful suggestions. Acta Paediatr. 2006;95(8):904–8.

81. Mathai S, Derraik JG, Cutfield WS, Dalziel SR, Harding JE, Biggs J, et al. Increased adiposity in adults born preterm and their children. PLoS One. 2013;8(11):e81840.

82. Blencowe H, Cousens S, Oestergaard MZ, Chou D, Moller AB, Narwal R, et al. National, regional, and worldwide estimates of preterm birth rates in the year 2010 with time trends since 1990 for selected countries: a systematic analysis and implications. Lancet. 2012;379(9832):2162–72.

83. Villar J, Papageorghiou AT, Knight HE, Gravett MG, Iams J, Waller SA, et al. The preterm birth syndrome: a prototype phenotypic classification. Am J Obstet Gynecol. 2012;206(2):119–23.

84. McElrath TF, Hecht JL, Dammann O, Boggess K, Onderdonk A, Markenson G, et al. Pregnancy disorders that lead to delivery before the 28th week of gestation: an epidemiologic approach to classification. Am J Epidemiol. 2008;168(9):980–9.

85. Sipola-Leppanen M, Kajantie E. Should we assess cardiovascular risk in young adults born preterm? Curr Opin Lipidol. 2015;26(4):282–7.

86. Jones EM SN, Gaye A, Laflamme P, Burton PR. Combined analysis of correlated data when data cannot be pooled. Stat. 2013;2.

87. Jones EM SN, Masca N, Wallace S, Murtagh MJ, Burton PR. DataSHIELD – shared individual-level analysis without sharing data: a biostatistical perspec-tive. Norwegian Journal of Epidemiology. 2012;21:231–9.

88. Reilly JJ, El-Hamdouchi A, Diouf A, Monyeki A, Somda SA. Determining the worldwide prevalence of obesity. Lancet. 2018;391(10132):1773–4.

